# SARS-CoV-2 N501Y introductions and transmissions in Switzerland from beginning of October 2020 to February 2021 – implementation of Swiss-wide diagnostic screening and whole genome sequencing

**DOI:** 10.1101/2021.02.11.21251589

**Authors:** Ana Rita Goncalves Cabecinhas, Tim Roloff, Madlen Stange, Claire Bertelli, Michael Huber, Alban Ramette, Chaoran Chen, Sarah Nadeau, Yannick Gerth, Sabine Yerly, Onya Opota, Trestan Pillonel, Tobias Schuster, Cesar M.J.A. Metzger, Jonas Sieber, Michael Bel, Nadia Wohlwend, Christian Baumann, Michel C. Koch, Pascal Bittel, Karoline Leuzinger, Myrta Brunner, Franziska Suter-Riniker, Livia Berlinger, Kirstine K. Søgaard, Christiane Beckmann, Christoph Noppen, Maurice Redondo, Ingrid Steffen, Helena M.B. Seth-Smith, Alfredo Mari, Reto Lienhard, Martin Risch, Oliver Nolte, Isabella Eckerle, Gladys Martinetti Lucchini, Emma B. Hodcroft, Richard A. Neher, Tanja Stadler, Hans H. Hirsch, Stephen L. Leib, Lorenz Risch, Laurent Kaiser, Alexandra Trkola, Gilbert Greub, Adrian Egli

## Abstract

The rapid spread of the SARS-CoV-2 lineages B.1.1.7 (N501Y.V1) throughout the UK, B.1.351 (N501Y.V2) in South Africa, and P.1 (B.1.1.28.1; N501Y.V3) in Brazil has led to the definition of variants of concern (VoCs) and recommendations by the European Center for Disease Prevention and Control (ECDC) and World Health Organization (WHO) for lineage specific surveillance. In Switzerland, during the last weeks of December 2020, we established a nationwide screening protocol across multiple laboratories, focusing first on epidemiological definitions based on travel history and the S gene dropout in certain diagnostic systems. In January 2021, we validated and implemented an N501Y-specific PCR to rapidly screen for VoCs, which are then confirmed using amplicon sequencing or whole genome sequencing (WGS). A total of 3492 VoCs have been identified since the detection of the first Swiss case in October 2020, with 1370 being B1.1.7, 61 B.1.351, and none P.1. The remaining 2061 cases of VoCs have been described without further lineage specification. In this paper, we describe the nationwide coordination and implementation process across laboratories, public health institutions, and researchers, the first results of our N501Y-specific variant screening, and the phylogenetic analysis of all available WGS data in Switzerland, that together identified the early introduction events and subsequent community spreading of the VoCs.

## Introduction

Since December 2020, three emerging SARS-CoV-2 lineages - B.1.1.7 (N501Y.V1), B.1.351 (N501Y.V2), and P.1 (B.1.1.28.1; N501Y.V3) - have generated concern in public and scientific communities. All three lineages show a rapid spread and displacement of locally established SARS-CoV-2 lineages, in the United Kingdom (UK), South Africa (ZA), and Brazil (BR), respectively, where they were first detected ^1-8^. The B.1.1.7 and B.1.351 lineages have subsequently been reported in many countries around the globe, including Switzerland. Most recently the P.1 lineage, exhibiting the N501Y and E484K mutations, among others, was described in Brazil ^9-11^ and has also been found in Japan ^12^. It is hypothesized that the viral variants B.1.1.7, B.1.351, and P.1 are more transmissible compared to other circulating variants, due to a higher affinity towards the angiotensin-converting enzyme 2 (ACE2) receptor resulting from the N501Y mutation ^13^ and were defined as variants of concern (VoC). In the last week of December 2020, the B.1.1.7 lineage accounted for more than 25% of overall published genomes from the UK (according to the Global Initiative on Sharing Avian Influenza Data (GISAID) as of 19 January 2021), but it is estimated to account for up to 70% of transmission events in specific areas of the UK ^14^. Waste water screening in Switzerland suggests that the B.1.1.7 lineage was present in Switzerland in early December ^15^. In South Africa, no reliable prevalence data on the B.1.351 lineage is available, but published data suggests that this VoC is also spreading more rapidly^6,16^).

The first genome belonging to the B.1.1.7 lineage was detected in September 2020 in the UK (according to the GISAID database) and showed 17 lineage specific polymorphisms, eight of which are located in the 1273 amino acid spike glycoprotein (nucleotide position 21563 to 25384, ^17-19^ **Table S1**). The spike glycoprotein is crucial for viral infection of host cells and is an important target for neutralizing antibodies ^20^. Some of the B.1.1.7 polymorphisms may modulate the protein’s function, such as the N501Y mutation in the receptor binding domain, the HV 69-70 deletion, and the P681H mutation in the furin cleavage site ^21,22^. The HV 69-70 deletion at nucleotide position 21765-21770 of the SARS-CoV-2 genome results in a dropout of the spike glycoprotein (S) gene diagnostic target in some commercial PCR assays. Although the S gene dropout is not specific for the B.1.1.7 lineage, it may nevertheless be a good first approach to screen for B.1.1.7 variants ^23,24^. This HV 69-70 deletion in the spike glycoprotein might favour immune escape ^17^. The B.1.1.7 variant also carries several lineage specific mutations in the ORF8 gene (**Table S1**), which might also be associated with decreased host immunity against SARS-CoV-2. Indeed, the ORF8 protein disrupts antigen presentation and reduces the recognition and the elimination of virus-infected cells by cytotoxic T-cells ^25^.

The B.1.351 lineage was first detected in October 2020 in ZA (according to the GISAID database) and also shares the N501Y mutation, but has otherwise different lineage-determining polymorphisms (**Table S1**) and does not show a characteristic S gene dropout due to lack of the HV 69-70 deletion. Of particular concern is the spike glycoprotein E484K mutation, which has been shown to reduce binding affinities towards neutralizing antibodies ^6,26,27^. Some of the polymorphisms that the viral variants described here possess are also present in other SARS-CoV-2 lineages (**Table S2**) and hence raise the question about how viral variants and lineages evolve (parallelism or same ancestral strain) and what selective pressures are important at single patient and population levels. The origins of B.1.1.7 and B.1.351 remain speculative, but include mutations during chronic infection in immunosuppressed patients exposed to convalescent plasma or other therapies ^28^, or potential recombination events between different lineages. As SARS-CoV-2 whole genome sequencing (WGS) is not performed uniformly across the globe, there may be other, unsampled, lineages also showing similar features of selection. Some variants may have been selected in intensive mink farms, where large outbreaks have been documented, as well as common cross-species transmission from human to minks and back ^29^. The adaptations on VoCs may lead to a substantially higher case burden ^5^, potentially paving the way for additional waves of the pandemic, and continued challenge for healthcare systems across European countries. Therefore, rapid identification of the B.1.1.7, B.1.351, and P.1 lineages is very important, and should trigger intensified contact tracing, targeted public health interventions in affected geographical areas, and re-allocation of vaccination strategies to areas with increasing community transmission of the VoCs.

During December 2020, awareness of the B.1.1.7 and B.1.351 lineages and the epidemiological situations in the UK and ZA reached the public, while at the same time approximately 10,000 tourists from endemic areas arrived in Switzerland for ski holidays. In order to understand the spread of VoC and to adapt public health interventions accordingly, a multi-step screening concept was developed across diagnostic and research laboratories in collaboration with the Federal Office of Public Health (FOPH), Spiez Laboratory from the Federal Office for Civil Protection (FOCP), Coordination Commission of Clinical Microbiology of the Swiss Society of Microbiology (CCCM-SSM), and the National Reference Center for Emerging Viral Infections at the University Hospital Geneva. The concept first focused on the epidemiological risk definition, with travel history from the UK and ZA. Second, a microbiological risk definition used the S gene dropout as a potential indicator for the B.1.1.7 lineage. Third, N501Y-specific PCRs were established in several laboratories. Suspected cases were confirmed by amplicon sequencing or whole genome sequencing for accurate lineage determination. This screening strategy was rapidly and sequentially implemented within a few weeks through 21 diagnostic laboratories (as of 02.02.2021) across Switzerland with the goals of reducing and delaying introduction and community transmission events of the B.1.1.7 and B.1.351 lineages within Switzerland. In this article, we share our experience of a nationwide screening strategy, its implementation, and early results on the spread of the VoCs in Switzerland.

## Methods

### Ethical statement

This study was conducted in close collaboration with the FOPH and part of an epidemiological assessment (Communicable Diseases Legislation – Epidemics Act). In addition, the study was approved as a multi-center study by the leading ethical committee (Ethik Kommission Nordwest-und Zentralschweiz, EKNZ; Approval number 2019-01291).

### Development of a screening strategy

Due to the highly probable introduction of the B.1.1.7 and B.1.351 lineages into the Swiss population, the FOPH, Spiez Laboratory (within the FOCP), the CCCM-SSM, the National Reference Center for Emerging Viral Infections, and the diagnostic laboratories developed a pragmatic screening strategy for the VoC (**Figure 1)**. The goal was to use already established infrastructures and reporting systems. The concept was communicated to cantonal physicians and diagnostic laboratories via the FOPH and FOCP and on the website of the CCCM-SSM ^30^. Suspected and confirmed VoCs were reported to the FOPH and cantonal physicians, initiating extensive backward and forward contact tracing with the goal of rapidly interrupting transmission chains. The screening strategy was continuously adapted.

**Figure 1.**
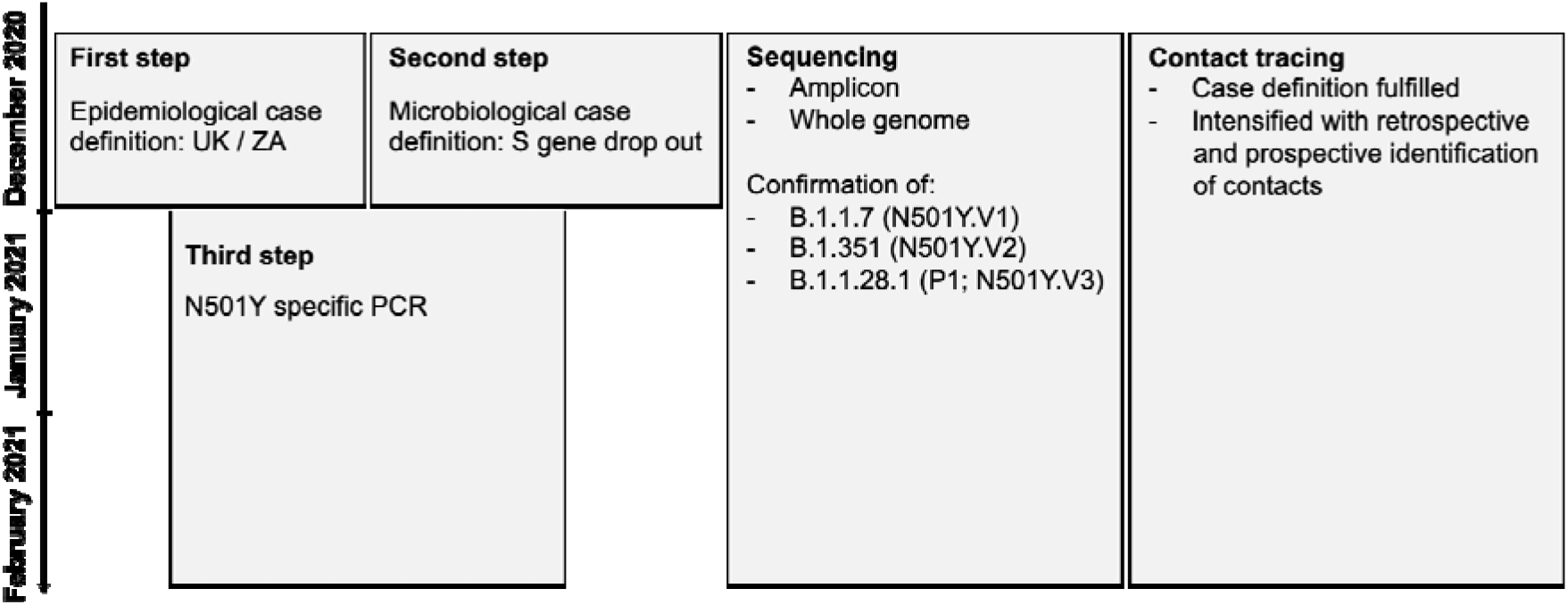
Diagnostic strategy to detect the B.1.1.7 and the B.1.351 in Switzerland. The flowchart shows the three step strategy with (i) initial epidemiological case definitions with travels from the UK or South Africa, (ii) the diagnostic evidence due to a S gene dropout and (iii) the final establishment of a N501Y-specific PCR. In all steps amplicon based and whole genome sequencing was used to determine and confirm the lineage allocation.

As a first step, an epidemiological case definition with recent travel history to the UK or ZA was used to identify potential carriers of the VoC. In mid-January 2021 this was expanded to BR. Both direct and indirect contacts of people travelling from these areas were considered. Patient travel history was recorded on mandatory FOPH reporting forms by clinical and laboratory institutions ^31^, as well as by cantonal physicians during contact tracing. In Switzerland, quarantine upon arrival was made mandatory for travellers from the UK and ZA from 28th December 2020; and from BR from 21st January 2021 ^32^.

As a second step, a microbiological case definition was used. LZM Risch AG using the TaqPath™ COVID-19 Combo Kit diagnostic assay (Thermo Fisher) noted a significant increase in S gene dropouts through November and December 2020. This multi-target PCR assay target sequences within the SARS-CoV-2 genes *ORF1ab, N* and *S*. A geographical distinction was observed: S gene dropouts were mainly noted in the eastern region of Switzerland, whereas other laboratories using the same assay in the western region of Switzerland noted an increase after a delay of four weeks. The S gene dropout is explained by a deletion at the positions 21765-21770 (HV 69-70) and is an indication for the B.1.1.7 lineage, as well as other non-VoC lineages. For this reason, and because initially variant-specific PCR assays were not yet available, the sequencing of all S gene dropouts was strongly recommended. A similar approach was developed by Danish ^33^ and Portuguese colleagues ^34^. However, during November and December, most of the S gene dropouts occurred in another emerging lineage, B.1.258, not showing the N501Y mutation, but featuring the HV 69-70 deletion and the N439K mutation ^35^.

During the Christmas holidays 2020, personnel and technical resources were limited, and focusing on these first two steps provided an initial screening program (from 22nd December,). In January 2021 the screening strategy was modified with a third step: several N501Y-specific PCR protocols were validated and established in laboratories throughout the country. All diagnostic laboratories in Switzerland were contacted on 15^th^ January 2021 via the FOCP to encourage and establish a N501Y-specific PCR. The CCCM-SSM published concomitantly additional recommendations on the society website (www.swissmicrobiology.ch). Since then, 21 laboratories have started to validate and implement a N501Y-specific PCR (as of 02.02.2021).

Various laboratories have used different PCR approaches to increase the pre-test probability of the identification of VoC. **Table S3** summarizes the different PCR approaches used by centers until 20th of January (**Table S3**). Most centers used the commercial assay SARS

Spike N501Y (53-0780-96; TIB MOLBIOL, Germany). In addition to the N501Y-specific PCR, at the University Hospital Lausanne, *ORF8* PCR/sequencing was used for the initial 12 samples received, in order to rapidly obtain results based on the presence/absence of the B.1.1.7 specific mutations C27972T, G28048T and A28111G ^17^, while waiting for the results of whole genome sequencing and the implementation of the S dropout and N501Y-specific PCR.

### Included samples for sequencing and reporting

The initial identified samples, from 22nd December 2020, were strongly biased towards the epidemiological and microbiological case definition (S gene dropout). From the beginning of January 2021, an increasing number of laboratories have joined the incentive and implemented N501Y-specific protocols. Meanwhile, older samples collected from September to December 2020 have also been sequenced. All VoC were reported to the FOPH via an electronic reporting form.

### Sanger sequencing protocols

For the sake of rapidity, amplicon-based sequencing, focusing on the S gene, was established at the National Reference Center for Emerging Viral Infections (HUG, Virology Laboratory) and implemented by other laboratories. The detailed protocols are available online ^36^. In addition, amplicon-based sequencing focusing on the *ORF8* gene was performed at the Institute of Microbiology of the University Hospital Lausanne. Briefly, specific primers were used to generate an amplicon for Sanger sequencing. All sequences were then compared to available sequences on GISAID.

### Whole genome sequencing protocols

For this study whole genome sequencing data was produced using Illumina and Oxford Nanopore Technologies (ONT, Oxford, UK) sequencing. SARS-CoV-2 genomes were generally amplified following the amplicon sequencing strategy of the ARTIC protocol (https://artic.network/ncov-2019) with V.1 or V.3 primers and 150 nucleotide paired-end sequenced, on an Illumina platform e.g.^37,38^. Most laboratories used Illumina based library preparations (NexteraXT or Nextera Flex). At the University Hospital Lausanne, libraries were prepared using the CleanPlex 5 SARS-CoV-2 Panel (Paragon Genomics). Further technical details on different sequencing protocols have been published 37.

A typical Nanopore sequencing library consisted of the pooling of PCR amplicons generated according to the ARTIC v3 protocol (https://artic.network/ncov-2019), which generates 400 bp amplicons that overlap by approximately 20 bp. Library preparation was performed with SQK-LSK109 (ONT) according to the ONT “PCR tiling of COVID-19 virus” (version: PTC_9096_v109_revE_06Feb2020, last update: 26/03/2020). Reagents, quality control and flow cell preparation were as described previously ^39,40^. ONT sequencing was performed on a GridION X5 instrument (Oxford Nanopore Technologies) with real-time base calling enabled (*ont-guppy-for-gridion* v.4.2.3; fast base calling mode). Sequencing runs were terminated after production of at least 100,000 reads per sample. Bioinformatic analyses followed the workflow described (https://artic.network/ncov-2019/ncov2019-bioinformatics-sop.html) using artic version 1.1.3. Consensus sequences were generated using medaka (https://github.com/nanoporetech/medaka) and bcftools ^41^.

Each center used individual bioinformatic pipelines to check for sequencing quality and generate the consensus sequences details are shared in GISAID (e.g. ^37^; https://gitlab.com/RKIBioinformaticsPipelines/ncov_minipipe/). The consensus sequence data were either directly shared between diagnostic laboratories or via GISAID.

### Phylogenetic inference

Global sequences and metadata were downloaded from GISAID ^42,43^ (as of 2nd February 2021; 456,184 consensus sequences). Sequences with more than 10 percent Ns (94,271) and with incomplete dates (23,871) were removed. 342,970 sequences remained, which were joined with sequence data from the University Hospital Lausanne (N=59), which is not yet available on GISAID. The total dataset contained a total of 260 whole genomes from variants of concern with S:N501Y mutations from Switzerland (B.1.1.7, n=249; B.1.351, n=11)(**Table S4**). The latest collection date of a N501Y positive whole genome dates to 19th January 2021, the earliest to 28th November 2020. We inferred a time-calibrated phylogeny rooted to the first cases in Wuhan, China from December 2019, using a subset of global genomes and focal Swiss sequences. For sub-setting, we included 800 genomes evenly subsampled over canton (administrative subdivision), month and year for the focal area Switzerland; 20 genomes per country and month in Europe, and 5 genomes per country and month for the rest of the world (contextual samples) totalling 9,026 genomes, using the nextstrain software v.2.0.0.post1 (nextstrain.org) and augur v.10.3.8 ^44^. The resulting alignment of focal and contextual genomes was used to infer clusters with zero single nucleotide mutations (SNPs) using a custom python script (https://github.com/appliedmicrobiologyresearch). Identified clusters were investigated regarding cantonal origin of the sample as well as known travel history.

## Results

### SARS-CoV-2 case numbers and spatio-temporal distribution in Switzerland

The first cases of the B.1.1.7 lineage in Switzerland were detected in retrospectively examined and sequenced samples from UK travel returners dated back to mid-October 2020 in Geneva and Lausanne and end of November 2020 in Basel. The first cases of B.1.351 were discovered in December 2020 in Schwyz (GISAID ID Switzerland/SZ-ETHZ-410256/2020 and Switzerland/BS-UHB-11011756/2020) also in travel returnee from ZA.

The initial S gene dropout based screening showed a large geographical heterogeneity in terms of detected B.1.1.7 variants. Whereas in the eastern parts of Switzerland the S gene dropout based screening resulted only in about 1% of identified B.1.1.7 variants, in the western parts of Switzerland the percentage of B1.1.7 variants within the S gene dropouts was much higher e.g., at the University Hospital Lausanne, as many as 78.9% (101 of 128). Using our screening approach, a total of 3492 samples carrying VoC have been found across different geographical regions (cantons, administrative subdivisions) (**Table 1**; **Figure 2**) from 14th October 2020 to 5th February 2021. 1431 of 3492 (41%) could be confirmed by amplicon sequencing or whole genome sequencing, for which lineages were identified. 95.7% of these successfully sequenced genomes were assigned to the B.1.1.7 lineage. 4.3% were assigned to the B.1.351 lineage. No case of the P.1 lineage was detected.

**Table 1.**
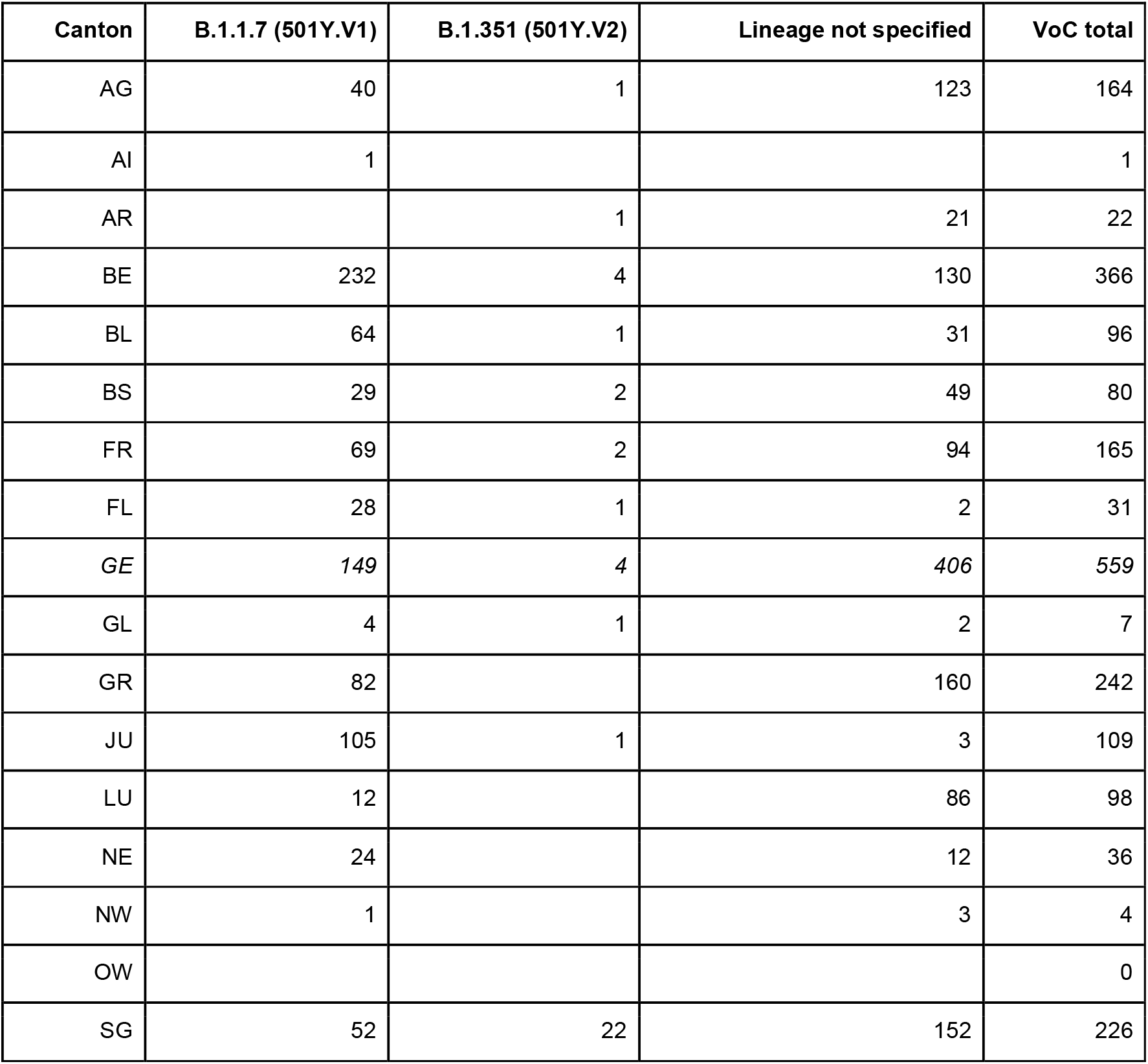
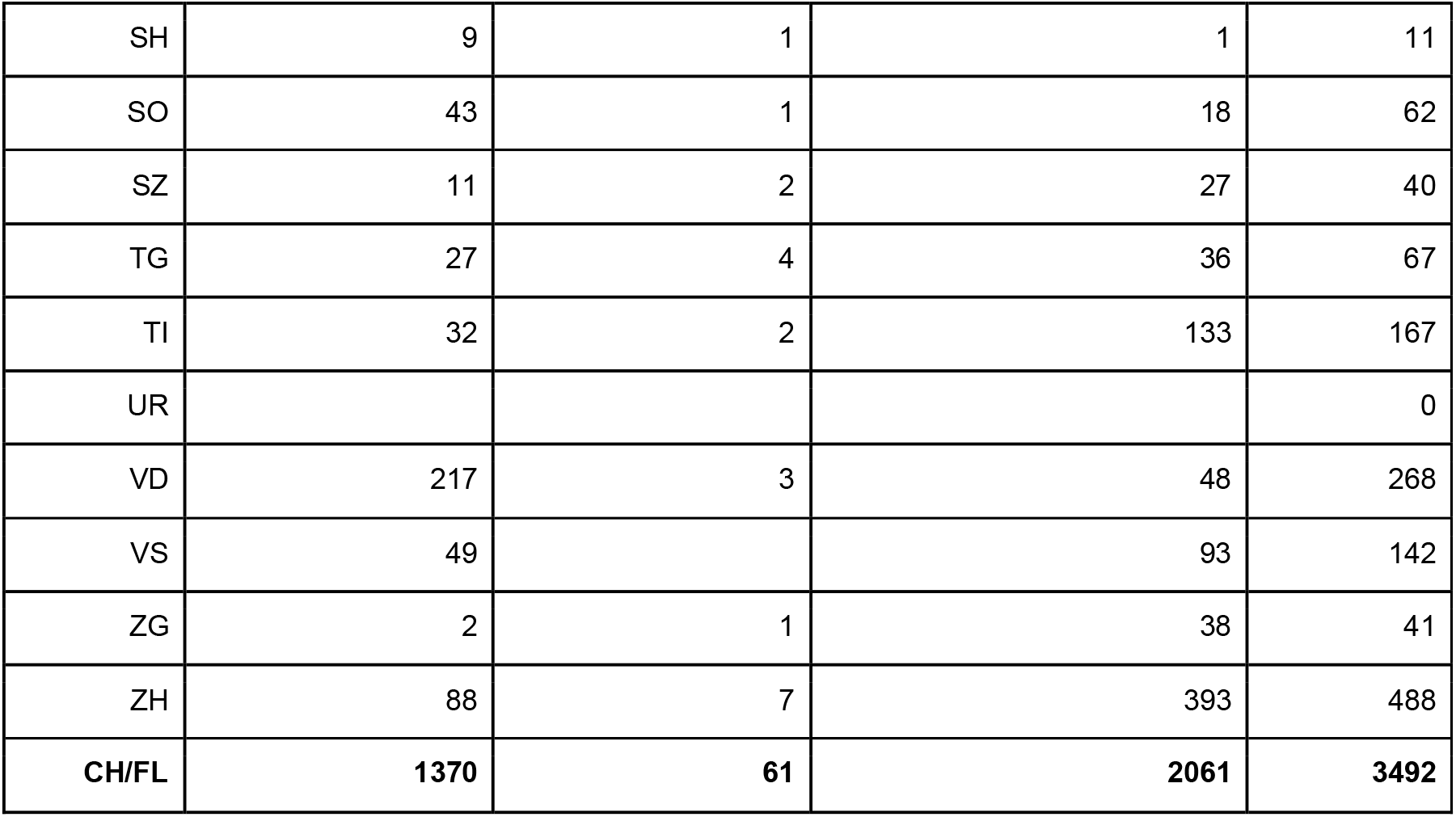
Absolute numbers of variants of concern (VoC) including cases of B.1.1.7 and B.1.351 in Switzerland and Principality of Liechtenstein. No case of P.1 was detected to 05.02.2021. Absolute numbers reflect a biased sample set due to the initial case definitions and biased distribution of diagnostic capacities. The numbers and distributions of lineages is likely biased due to delay in processing and different sequencing capacities.

**Figure 2.**
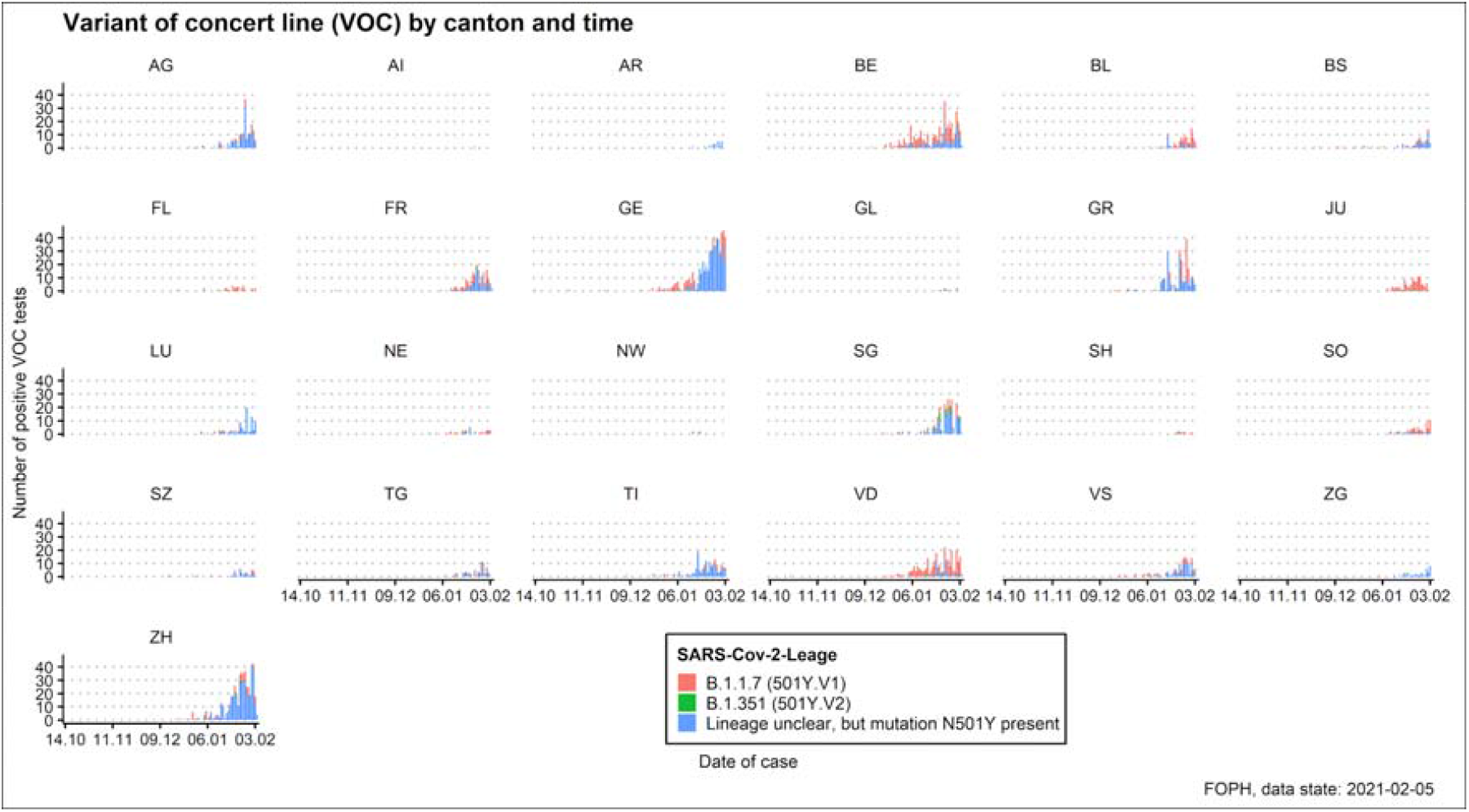
Distribution of absolute numbers as epidemiological curves according to canton and time. Absolute numbers reflect a biased sample set due to the initial case definitions, higher usage of antigen test in some regions, and distribution of diagnostic capacities. This does not reflect the prevalence of cases. Also the current amount of specific lineages is biased due to different sequencing capacities.

**Figure 3.**
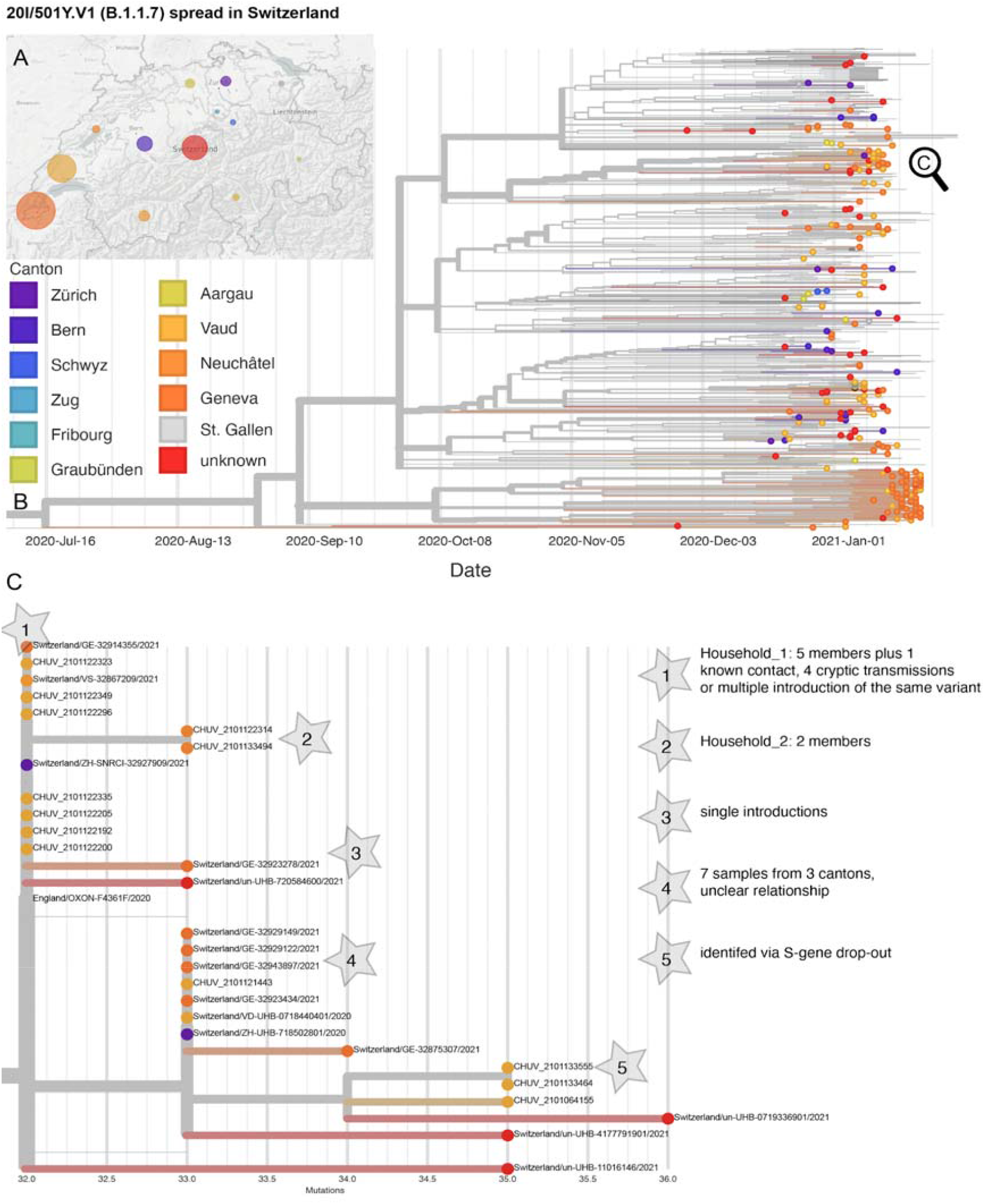
Phylogeny of sequenced B.1.1.7 cases in Switzerland. (A) Geographic distribution through Switzerland and (B) phylogenetic relationship of 249 genomes dating between 28th November, 2020 and 19th January 2021 from the Geneva University Hospitals (n=113), University Hospital Lausanne (n=59), University Hospital Basel (n=55), University of Bern (n=7), University of Zurich (n=12), and ETH Zurich (n=3). X-Axis scales to time. (C) Zoom into an exemplar constituting household transmissions, possible cryptic transmissions within a cluster, and single introductions, scale by mutations distance to the reference Wuhan/Hu1-1..

This initial screening was implemented in a few centers in the last two weeks of December 2020, during which time there was a selection bias due to the case definition and limited sequencing capacity over the Christmas holidays. Complementary to these cases, two additional datasets were analysed using whole genome sequencing: the first included 545 randomly selected SARS-CoV-2 samples from throughout Switzerland (Viollier AG, a large private laboratory) from 18th to 24th December 2020. Within this first dataset, no VoC was found. A second set focused on 1511 samples showing the S gene dropout (LMZ Dr. Risch, a large private laboratory) from 21st to 27th December 2020. From a total of 1511 PCR positive cases (15.1% positivity rate), 137 (9.1%) samples showed an S gene dropout. 79 of 137 S gene dropout samples were sequenced - and in 6/79 (8%) a B.1.1.7 lineage was found. This corresponds to 0.06% of the overall SARS-CoV-2 positive cases.

Since the second week of January 2021, increasing numbers of SARS-CoV-2 positive samples were analysed using an N501Y-specific PCR. However, at this stage our data does not allow the reliable determination of a Swiss-wide prevalence, as not all PCR positive cases are fully re-analysed with the N501Y-specific PCR. However, some laboratories re-analyse every SARS-CoV-2 positive case and thereby individual prevalence rates for VoCs could be determined for the last two weeks: The University Hospital Geneva reported 40% positivity for VoCs (25th to 31th of January); University of Zurich reported 13.3% (18th to 24th January), 20.2% (25th to 31th of January), and 28.4% (1st to 5th of February); Viollier AG reported 15% (25th to 31th of January) and 19.9% (1st to 5th of February); University Hospital Basel reported 29% (25th to 31th January) and 45% (1st to 4th of February); University of Bern reported 10.2% (25th to 31st January) and 30% (1st to 3rd February); Bioanalytica reported 6% (25th to 31st January) and 21.6% (1st to 5th February). LMZ Dr. Risch reported 18.5% (25th to 31st January) and 21% (1st to 3rd February). Within the upcoming weeks, we expect that a Swiss wide prevalence determination for VoC is established and reported. Of note, the detection rate has been going down in some centers and also overall, albeit slowly.

Some laboratories have reported the median age (with interquartile ranges) in years between patients with and without the N501Y variants. At the University Hospital Basel the median age of patients with N501Y positive was 34 years (IQR 12-47) whereas the median age of patients with N501Y negative was 38 years (27-54); At the University of Bern the media age was 33 years (IQR 20-51; N501Y positive) vs. 44 years (IQR 29-60; N501Y negative); at Bioanalytica the median age was 43 years (IQR 29-53; N501Y positive) vs. 48 years (IQR 32-67; N501Y negative); and at Viollier AG the median age was 41 years (IQR 26-54; N501Y positive) vs. 41 years (IQR 25-57; N501Y negative). Also this data may be biased due to the fact that certain laboratories may receive samples more predominantly from paediatric physicians or hospitals.

### Phylogenetic relatedness of first cases

A total of 260 N501Y-carrying (B.1.1.7 n=249, and B.1.351 n=11) high quality genomes from Switzerland were available for phylogenetic analysis. For 58 cases (known for University Hospitals Basel and Lausanne) a travel history to an endemic country or known contact to a traveller was available, however, for most cases the risk exposure was not available. For 213 cases the canton of residence was known. Using a 0 SNP threshold, we infer 9 out of the 11 B.1.351 cases to be single introduction events, two cases (from BS) are genetically (0 SNPs) and epidemiologically linked and trace back to a ZA travel returner and a transmission to a family member (**Figure S3**). The phylogenetic analysis of B.1.1.7 cases shows at least 116 single introductions into 11 cantons (**Figure 2A**), 106 without immediate links (0 SNP distance) to other genomes in the sub-sampled global dataset, 11 of which were known risk contacts or travellers (**Figure 2B**). Ten further single introductions had genetic links to genomes from UK samples, two of which had known travel history or risk contact. We identified 45 clusters (0 SNP distance) comprising 133 (range 2-10) genomes. 18 clusters contain samples with known travel links to the UK or risk contacts. Of interest, ten of these clusters contained cases from different cantons – suggesting outside of household transmission (**Figure 2C**).

## Discussion

The epidemiological situation with the SARS-CoV-2 lineages B.1.1.7 and B.1.351 in the UK and ZA resulted in the definition of so called variants of concern (VoC). The European Center for Disease Prevention and Control (ECDC) and the World Health Organization (WHO) strongly recommend identifying the viral lineages in order to monitor the distribution of VoCs, using sequencing for surveillance ^45^. In Switzerland, we started a targeted screening program for VoC in December 2020, and are currently developing an unbiased sequencing-based surveillance program for new variants. Interestingly, we found the first case of the B.1.1.7 lineage from October 2020, by sequencing archived sample collections However, in two large sets of 549 and 1511 SARS-CoV-2 samples from mid-to late-December 2020, we detected only sporadic cases of VoCs, suggesting that until then introductions and spread were not extensive. We calculate that the overall prevalence in Switzerland was less than 1% until the end of December 2020. Since then, our screening strategy showed a continuous increase of absolute case numbers, starting to ramp up in January 2021. This followed the Christmas holidays with thousands of ski tourists from endemic areas sojourning in Swiss ski resorts. During the first wave of the pandemic in 2020, skiing and associated activities resulted in an European wide spread of specific mutants linked to the alpine village Ischgl in Austria ^46,47^. Similar, we have detected cases linked to a potential super spreading event in the ski resort Wengen in December 2020, as reported in public news articles. Due to the concerns of high transmissibility of VoC, travellers from the UK and ZA were added to the quarantine list during the last week of December 2020. The comparison of our WGS data with a global dataset suggests direct links to the UK and indicates that a substantial number of the cases in January 2021 were due to individual introduction events from the UK. Unfortunately, information on links to UK travel is incomplete. Our experience suggests that detailed epidemiological data should be asked for and made available (such as travel history, contact to other infected people etc). In the future, more resources should be dedicated to this purpose. The identification of identical genomes from samples collected in distant regions of the country might indicate that cryptic transmission events are already happening as only a few cases of household transmission were documented. However, they could also be due to multiple introductions of identical variants from the UK to Switzerland and is a possibility that we cannot rule out.

Our sampling strategy focusing first on the epidemiological risk and a microbiological case definition including the S gene dropout, and the initial lack of diagnostic capabilities to confirm the VoC introduced a strong selection bias of samples. Thus our findings should be interpreted with care. Currently, our data does not allow us to properly determine the prevalence of VoCs in Switzerland. However, some laboratories have established a workflow including N501Y-specific PCR for all SARS-CoV-2 positive cases -this allowed us to monitor the increase in prevalence across tested samples in these individual laboratories. Available prevalence data for Switzerland is continuously visualized (https://ibz-shiny.ethz.ch/covidDashboard/variant-plot/index.html and https://ispmbern.github.io/covid-19/variants). Nevertheless, our pragmatic approach indicates that prior to the Christmas holidays, the VoC distribution in Switzerland was low (<1%) and now has reached within 8 weeks most likely rates of 20-45%. Our phylogenetic analysis provides important evidence that community transmission started to increase after New Year and continuously accelerated in mid-January 2021.

VoC identification based on the S gene dropout is not specific and sensitive enough to identify VoC lineages (see **Table S1**). Our WGS data of samples collected in December 2020 showed that most of the S gene dropout samples were due to the B.1.258 lineage, at least in eastern Switzerland. Due to this geographical distribution pattern, the curious phenomenon emerged that in western Switzerland the S dropout screen was an efficient approach to detect the B.1.1.7. lineage, whereas in eastern Switzerland it was confounded by other more prevalent lineages. For this reason, the pre-test probability for the B.1.1.7 lineage using the S dropout was different based on the local epidemiology.

The B.1.351 and P.1 lineages cannot be identified based on the S gene dropout. VoC can be confirmed using either amplicon based sequencing of the S gene or whole genome sequencing. Both sequencing approaches show specific advantages and disadvantages in terms of speed, costs, and resolution for molecular epidemiological studies. These aspects have to be carefully evaluated when establishing a screening program. Mixed usage may allow the strength of both methods. In order to efficiently select samples for subsequent sequencing, we have implemented a N501Y-specific PCR in many laboratories throughout Switzerland. The challenge was, as most diagnostic institutions were focusing on high throughput testing using fully automated robotic systems, to establish again a manual method including separate RNA extraction. It took several weeks to establish the workflows in larger laboratories and to implement new variables for reporting. Similar to the UK, our data shows an increase of positivity rates across time and the B.1.1.7 lineage displaces other circulating strains.

Surveillance of these and other VoC may become more important with new selective pressures such as therapeutics and vaccination. The Swiss model to monitor in a first phase with epidemiological and microbiological case definitions and in a second phase, as soon as it was available, with a specific PCR, allowed to rapidly screen isolates and identify the N501Y mutation as a surrogate marker for a potentially more transmissible variant. The subsequent confirmation with sequencing provides an efficient way to rapidly identify certain VoCs. We feel it is strongly recommended to further sequence the VoCs and not stop at the identification of the N501Y mutations. Lineage or also whole genome resolution provides highly valuable information for public health management ^48^. The development of our screening system, with all attached pre-to post-analytical aspects of diagnostics, may be valuable in the search for future upcoming variants such as vaccine escape mutants. However, it is clearly time for nations to seriously consider implementing national surveillance programs with an unbiased sequencing approach, incorporating sustainable elements for other key pathogens and potential future pandemics ^49^.

## Data Availability

Genomic data is semi-publicly available on GISAID.org. Summary statistics were made available directly by the Federal Office of Public Health (FOPH) Switzerland.

## Supplementary Tables

**Table S1.**
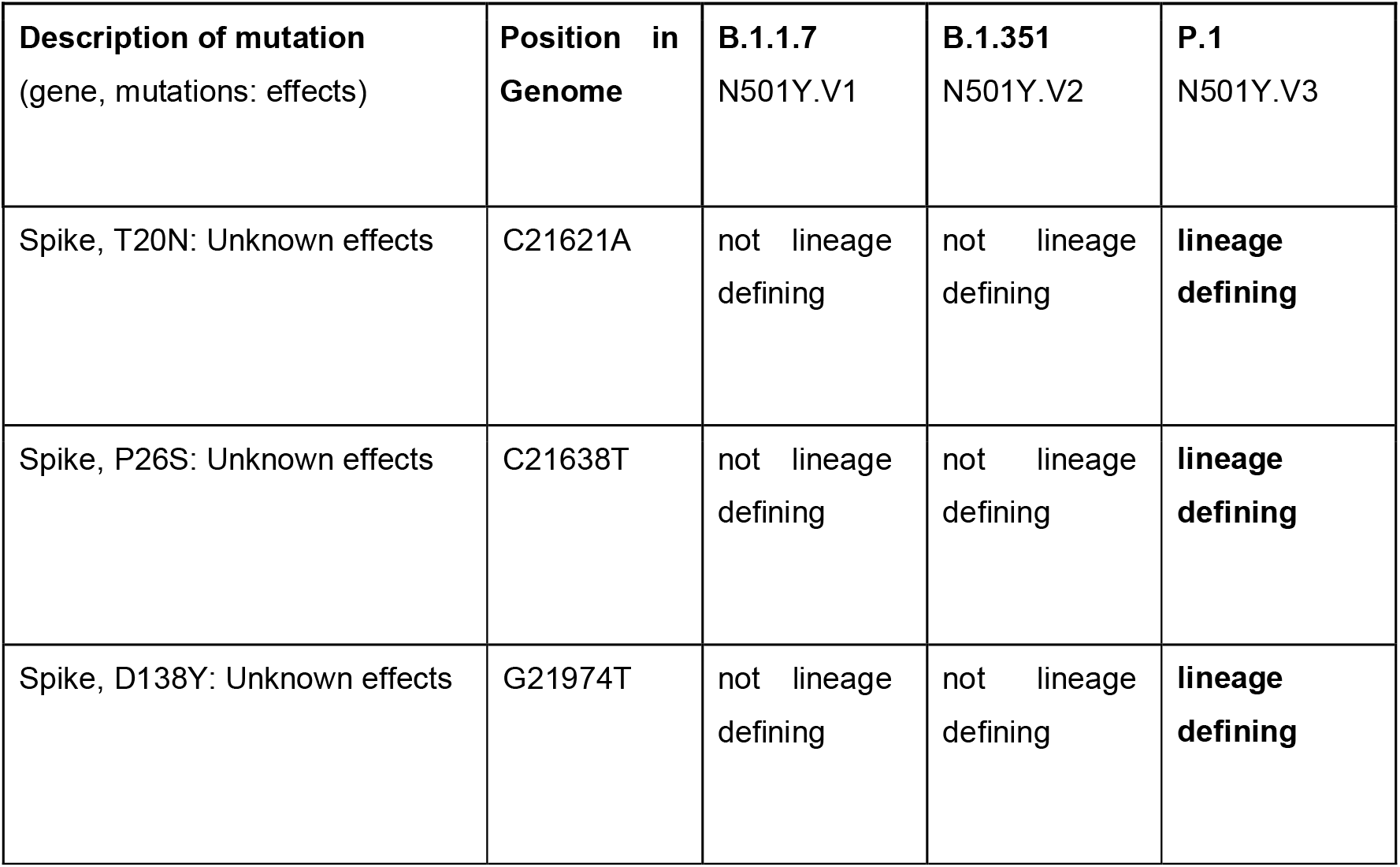
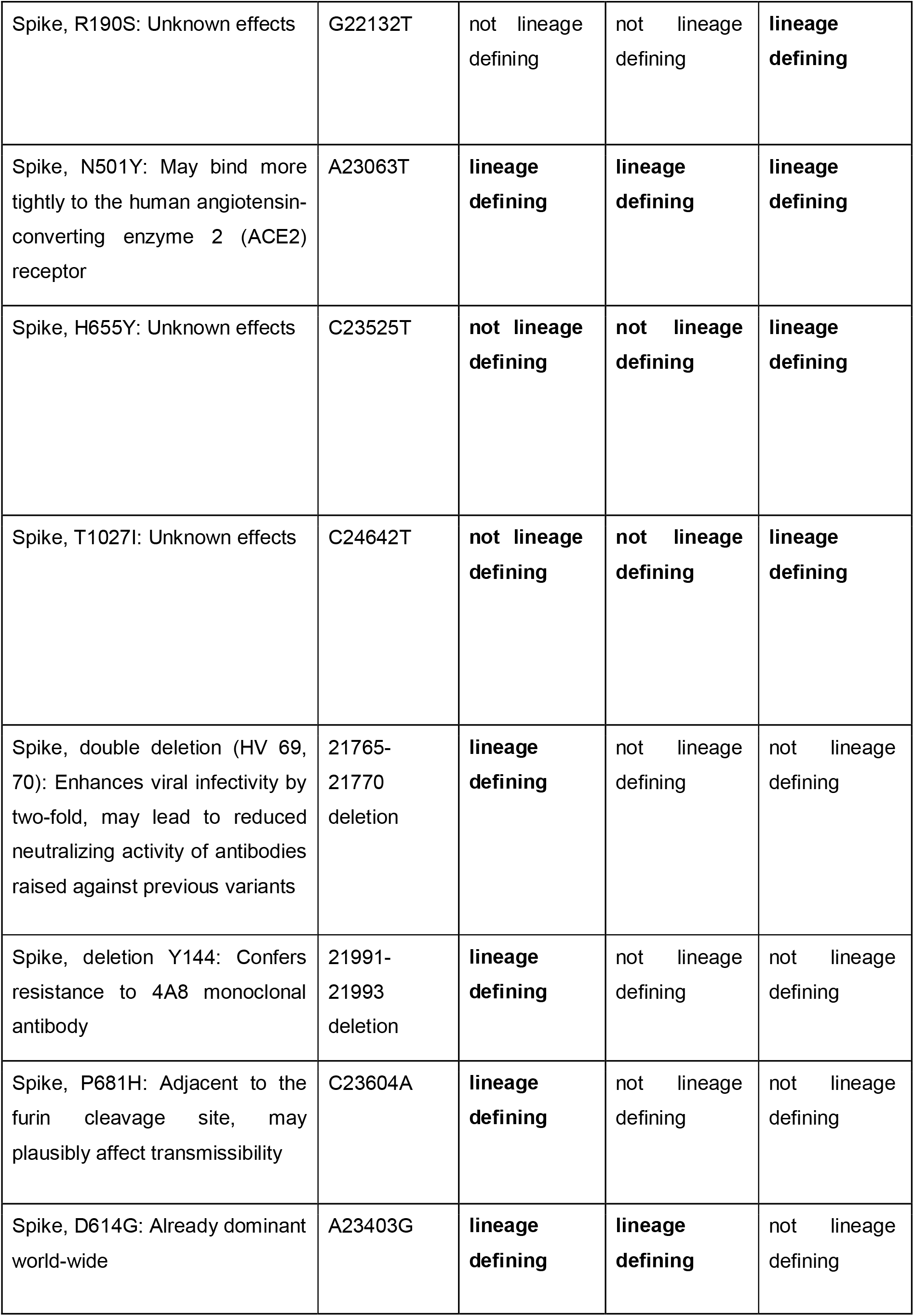
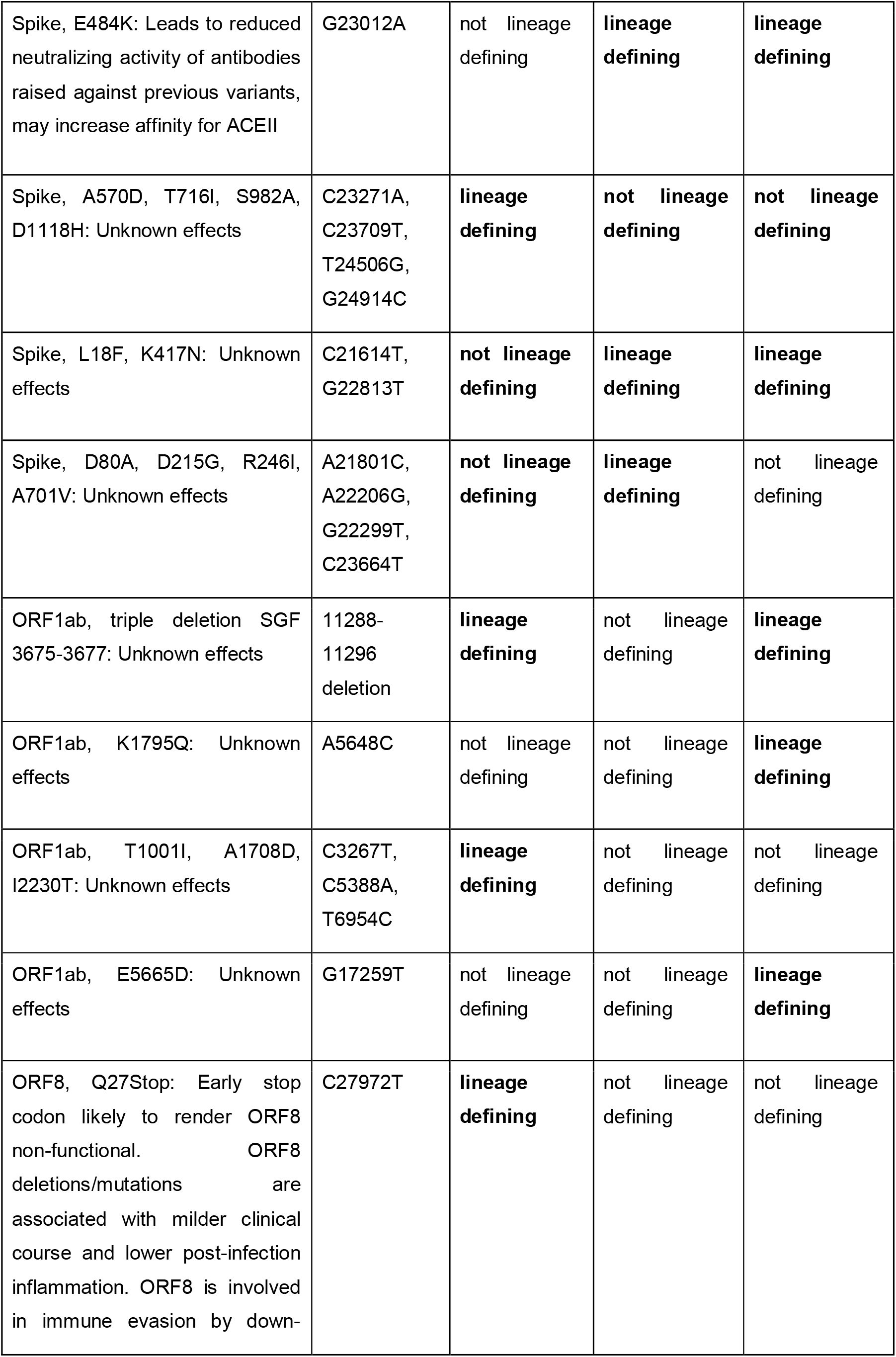
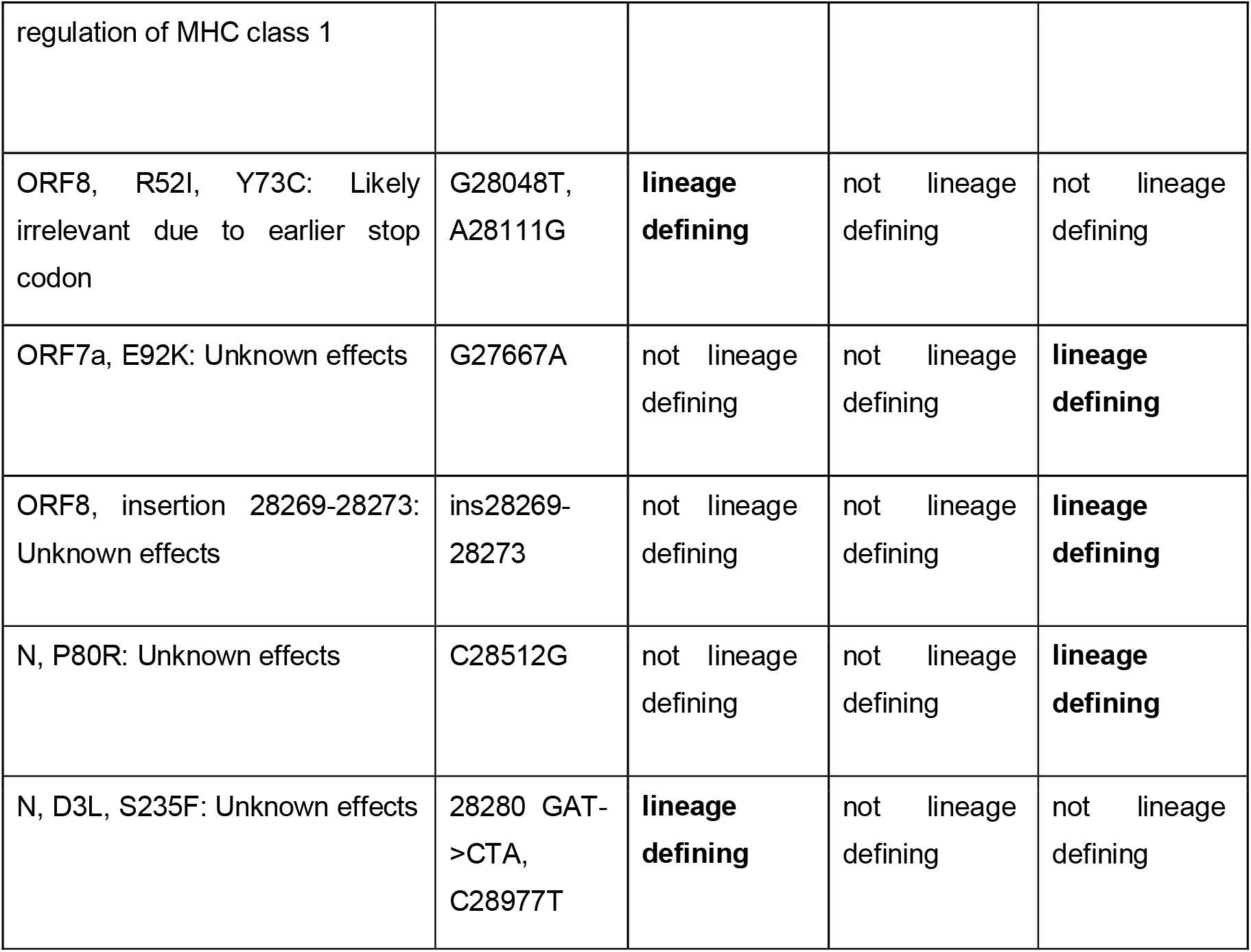
Mutations of the new SARS-CoV-2 lineages B.1.1.7, B.1.351 and P.1 (B.1.1.28.1). Data from GISAID and Covariants.org/shared-mutations. Full SARS-CoV-2 genome of Wuhan-Hu-1 is available (https://www.ncbi.nlm.nih.gov/nuccore/MN908947).

**Table S2.**
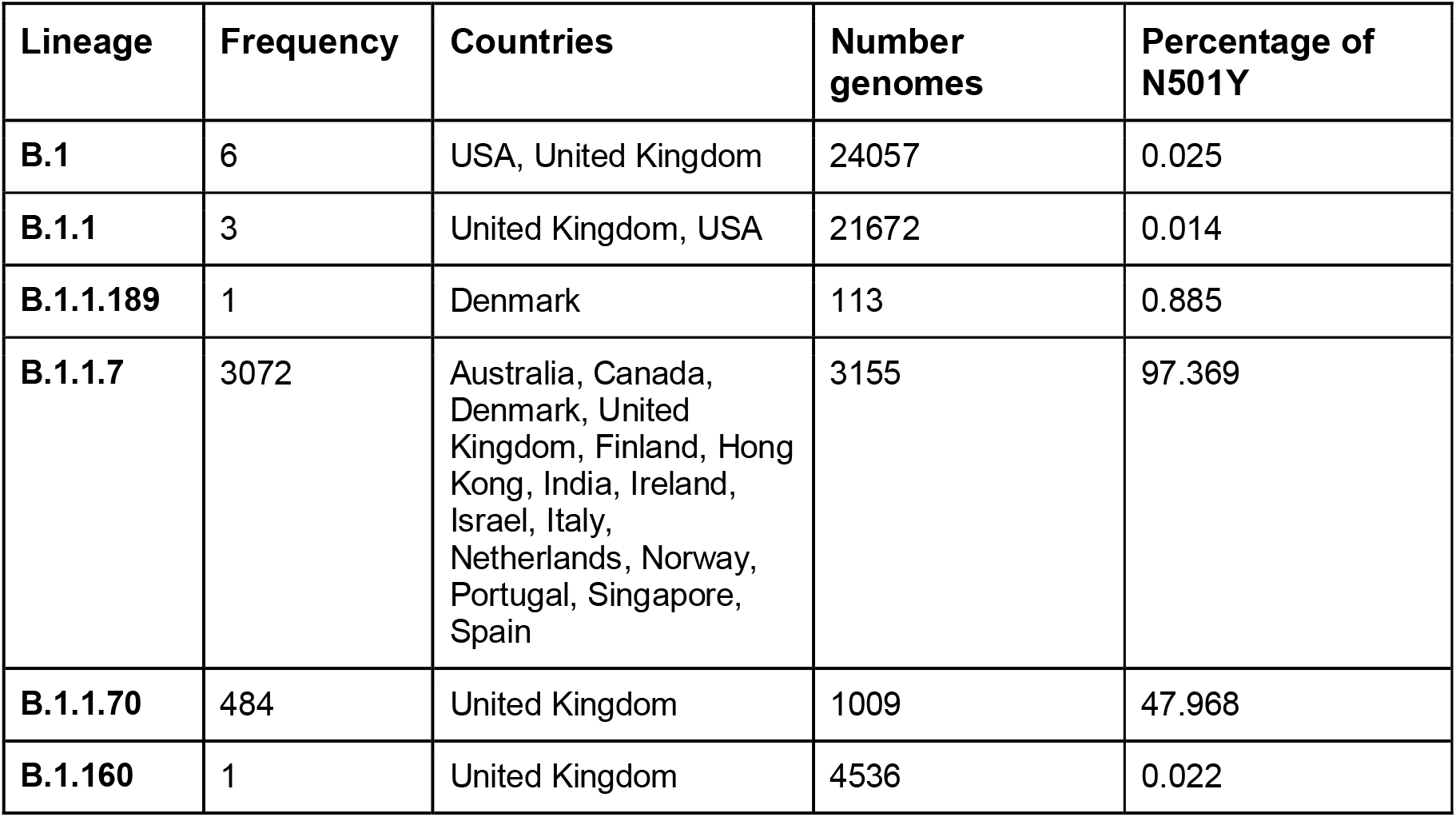
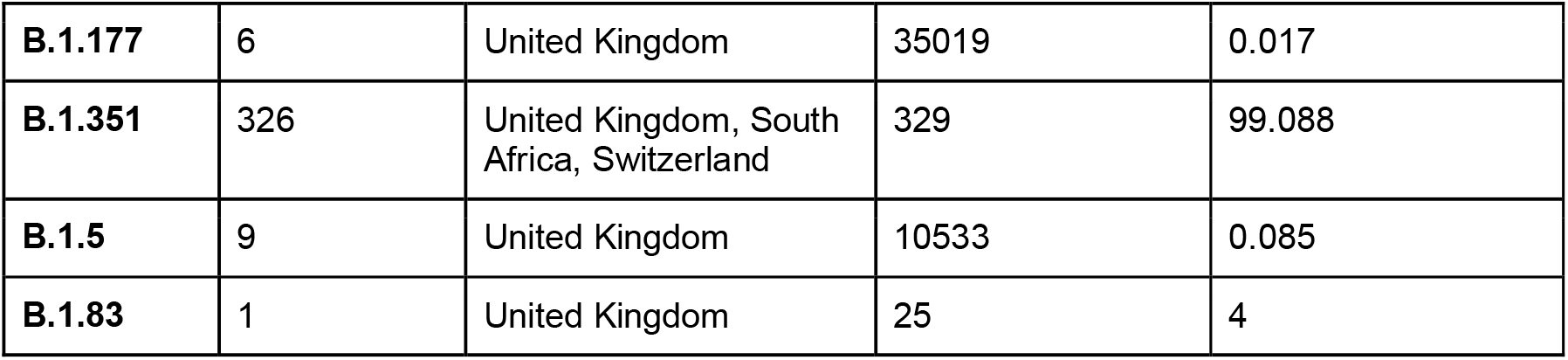
N501Y mutations across different viral lineages since September 2020. Based on GISAID database (access 15.01.2021).

**Table S3.**
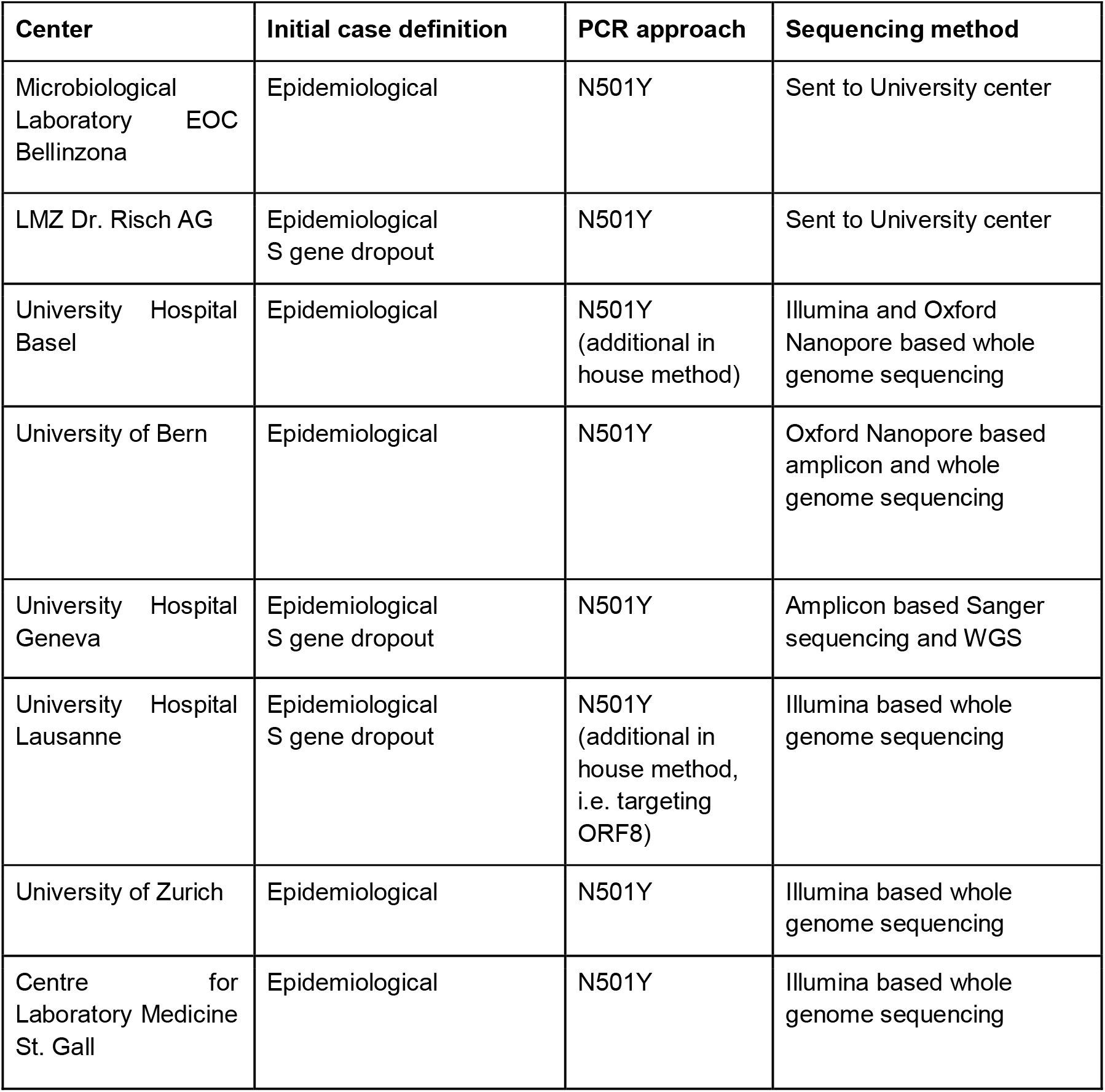
Screening methods used by different diagnostic laboratories as of 20th January 2021. N501Y screening assay was most commonly used from TIB MOLBIOL (Germany), but also in-house developed assays were used. University centers offering whole genome sequencing included the University Hospital Basel (Division of Clinical Bacteriology and Mycology), University of Bern (Institute for Infectious Diseases), University Hospital Geneva (Virology Laboratory), University Hospital Lausanne (Institute of Microbiology), and the University of Zurich (Institute of Medical Virology). In addition, ETHZ (D-BSSE core facility) sequenced samples.

**Table S4.**
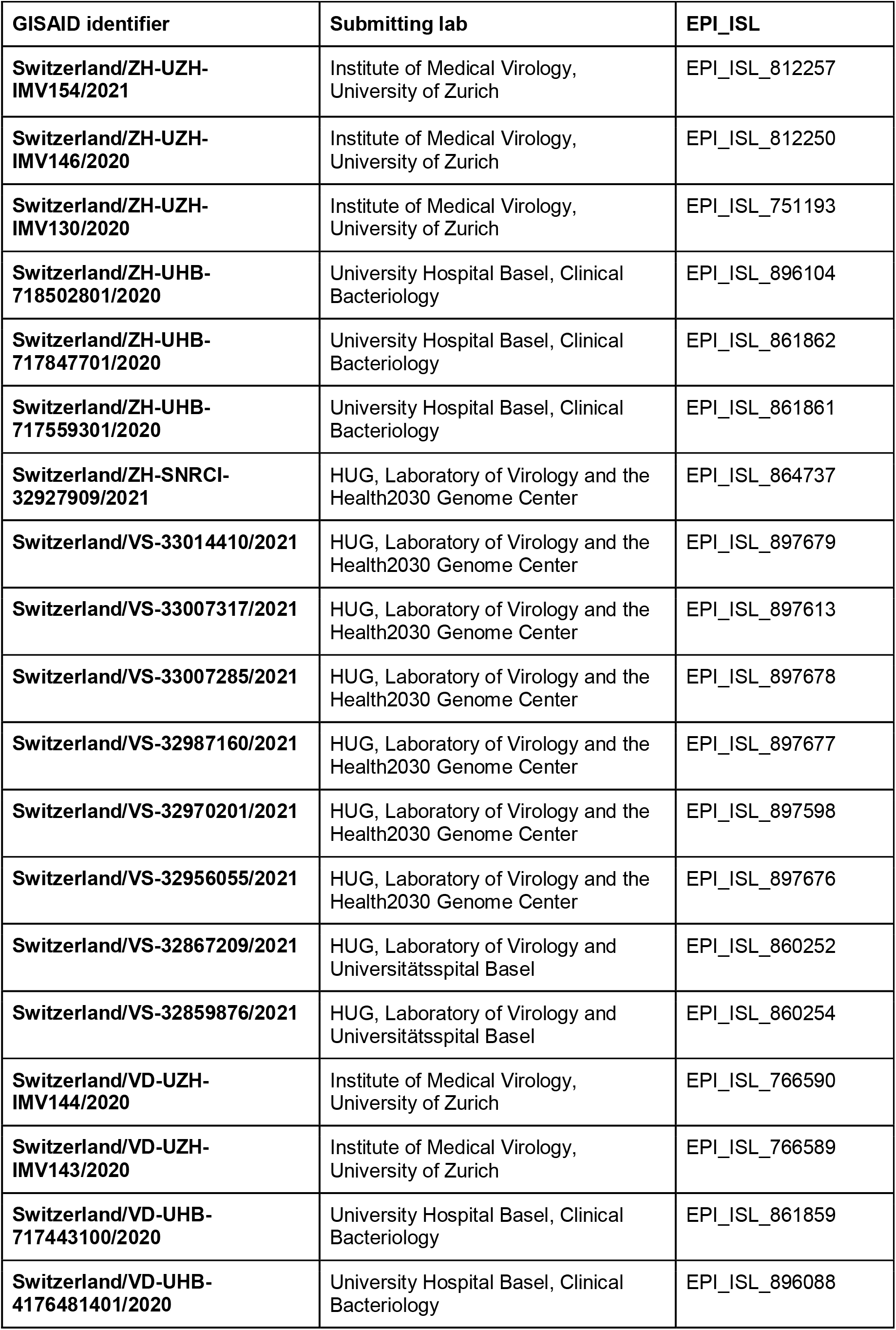
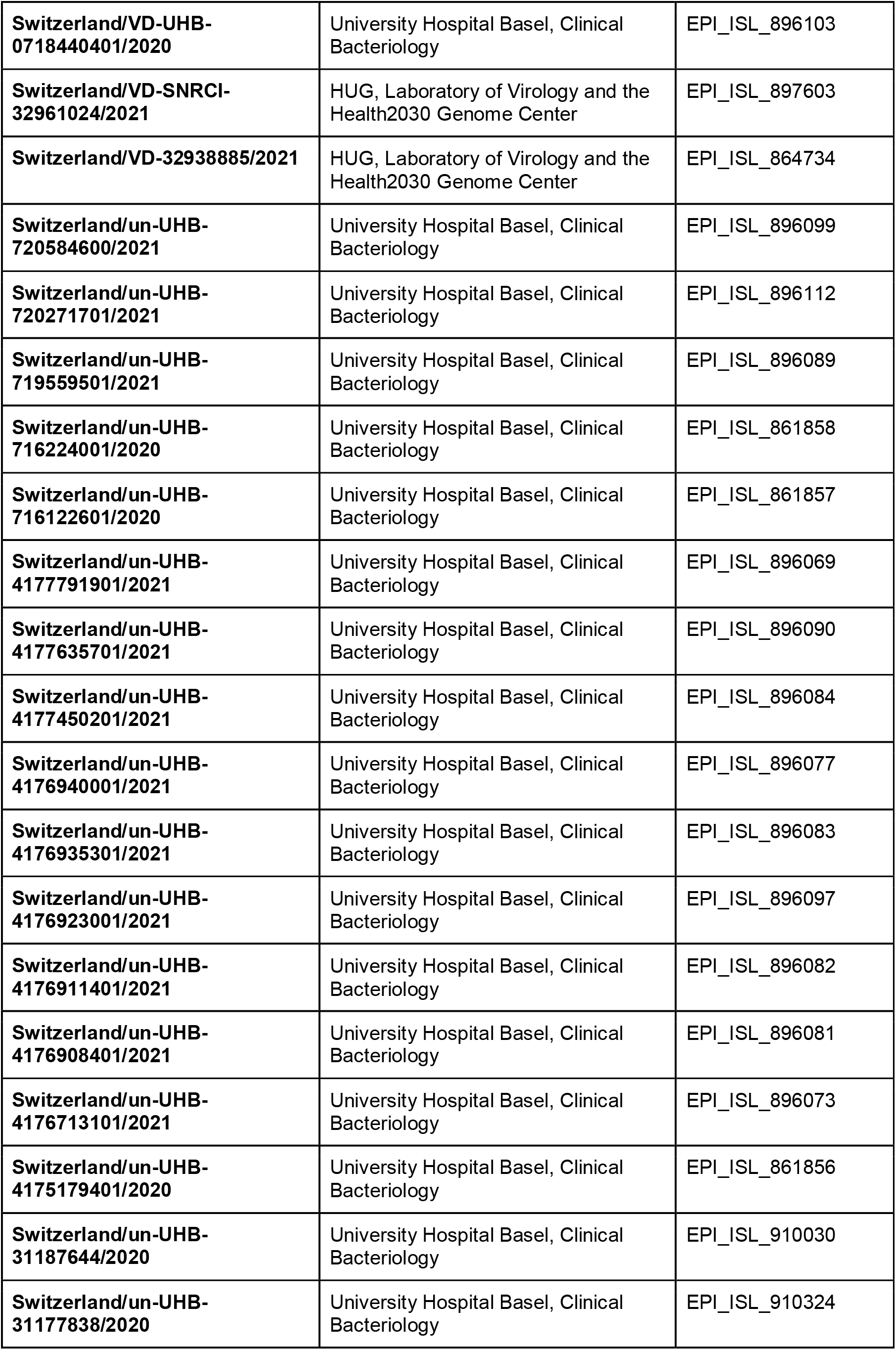
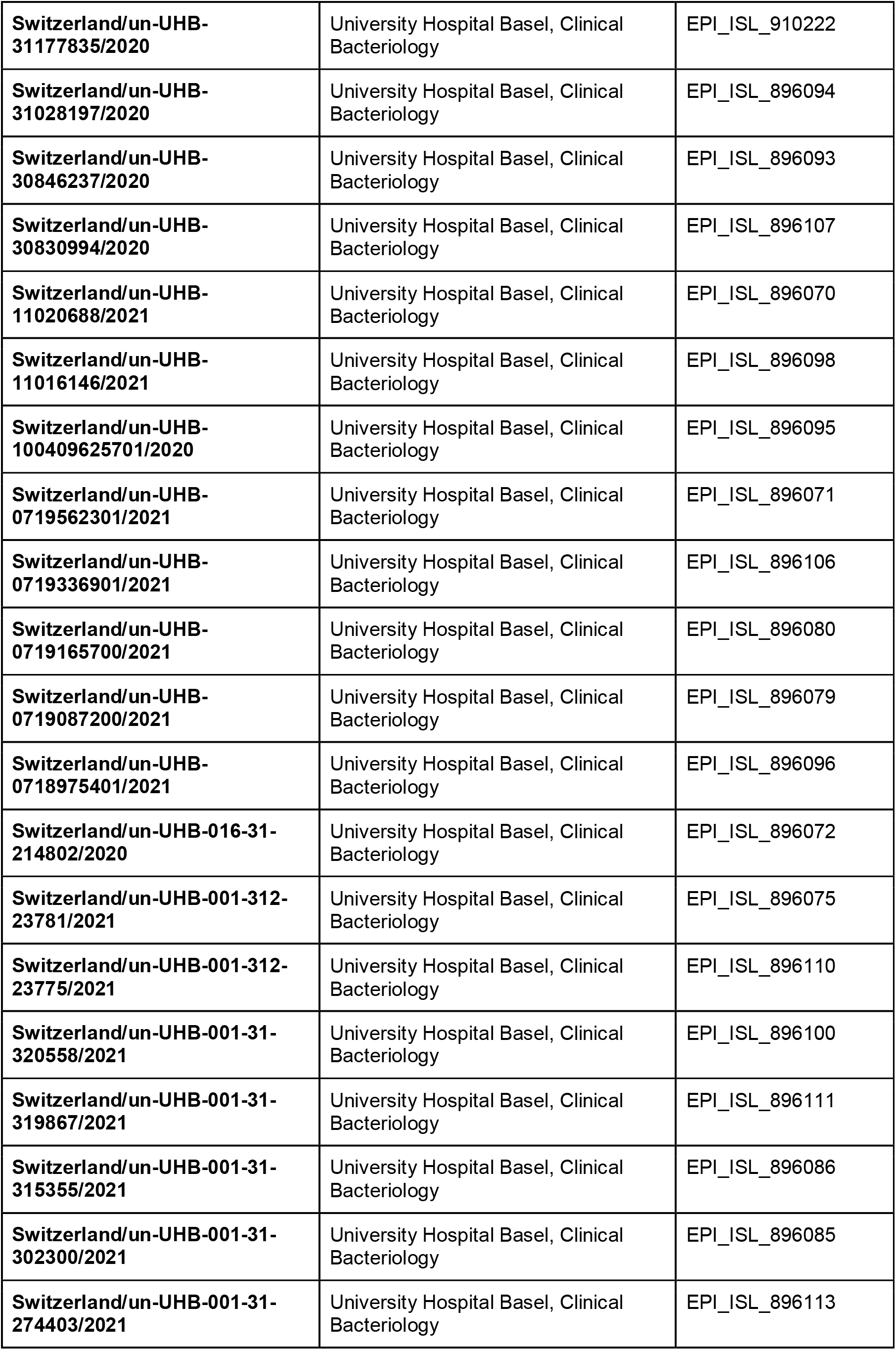
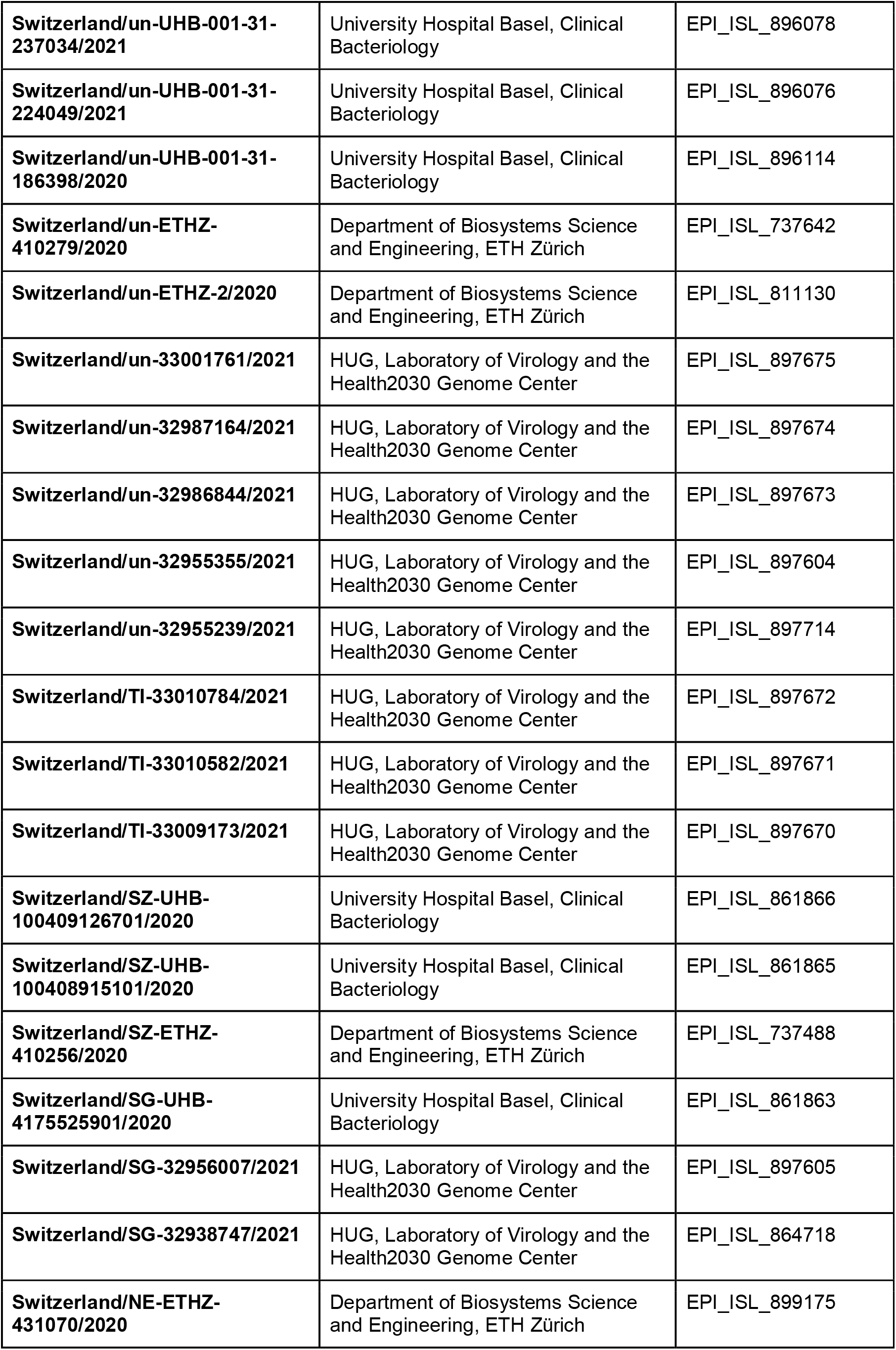
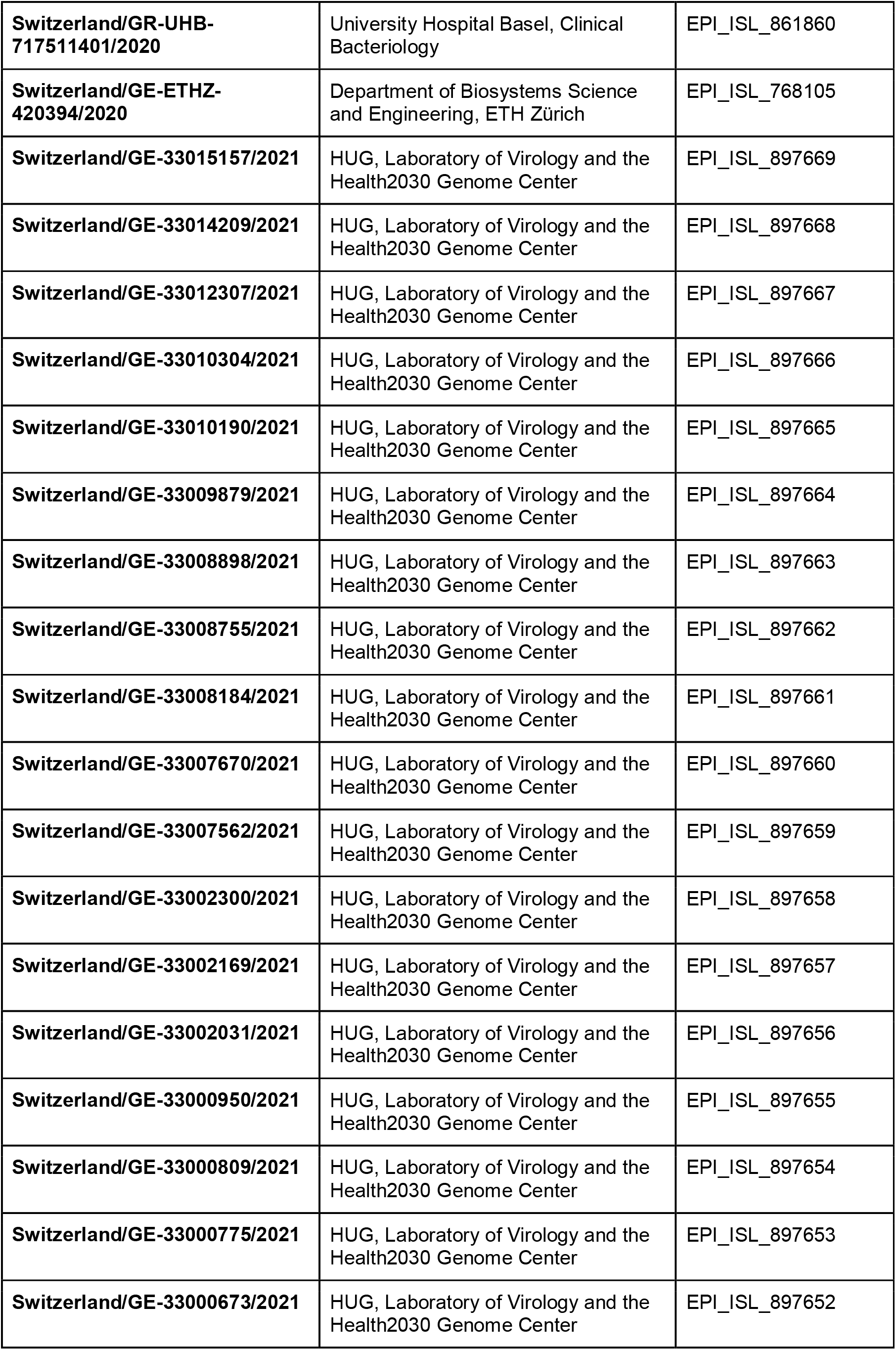
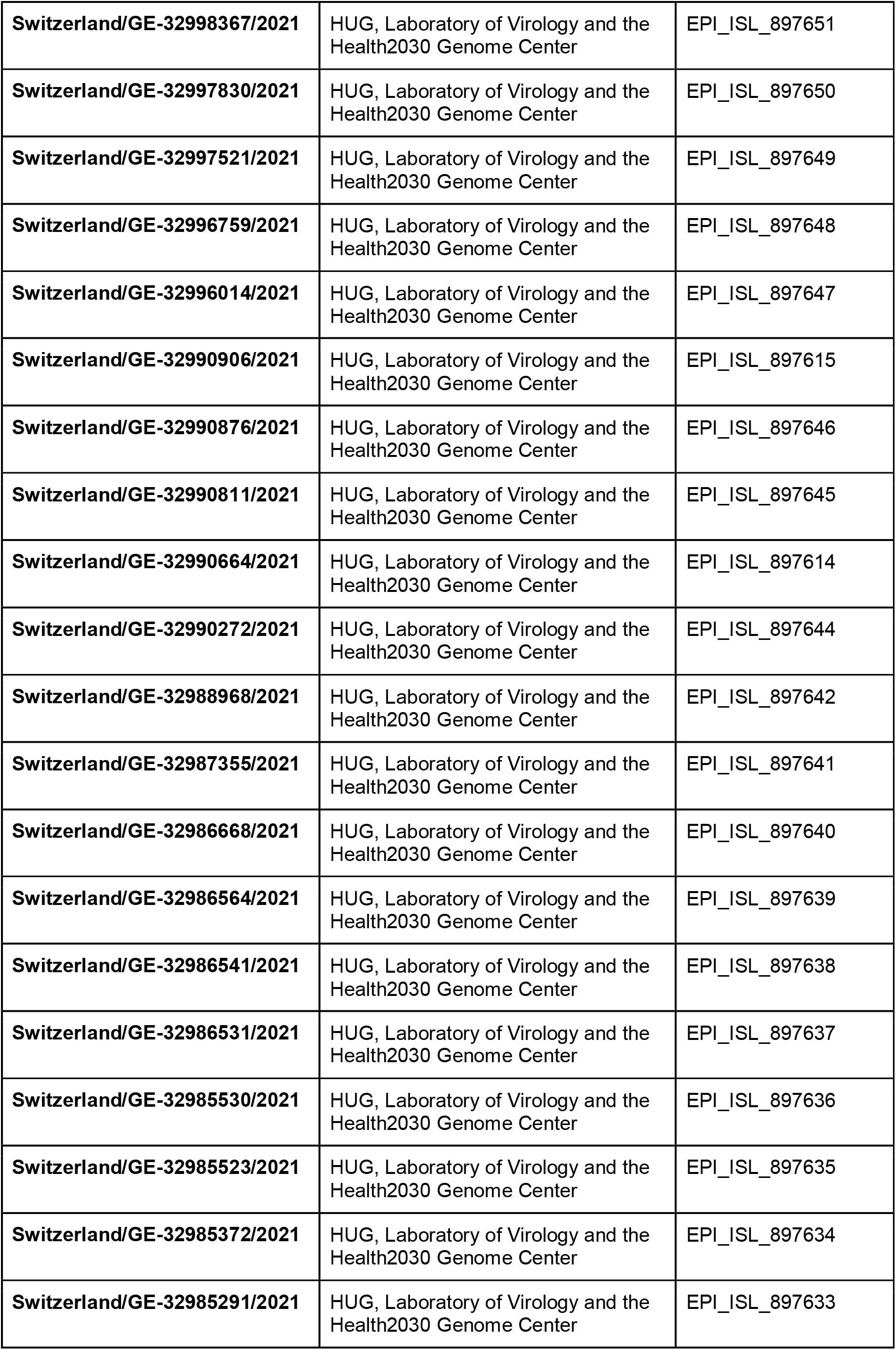
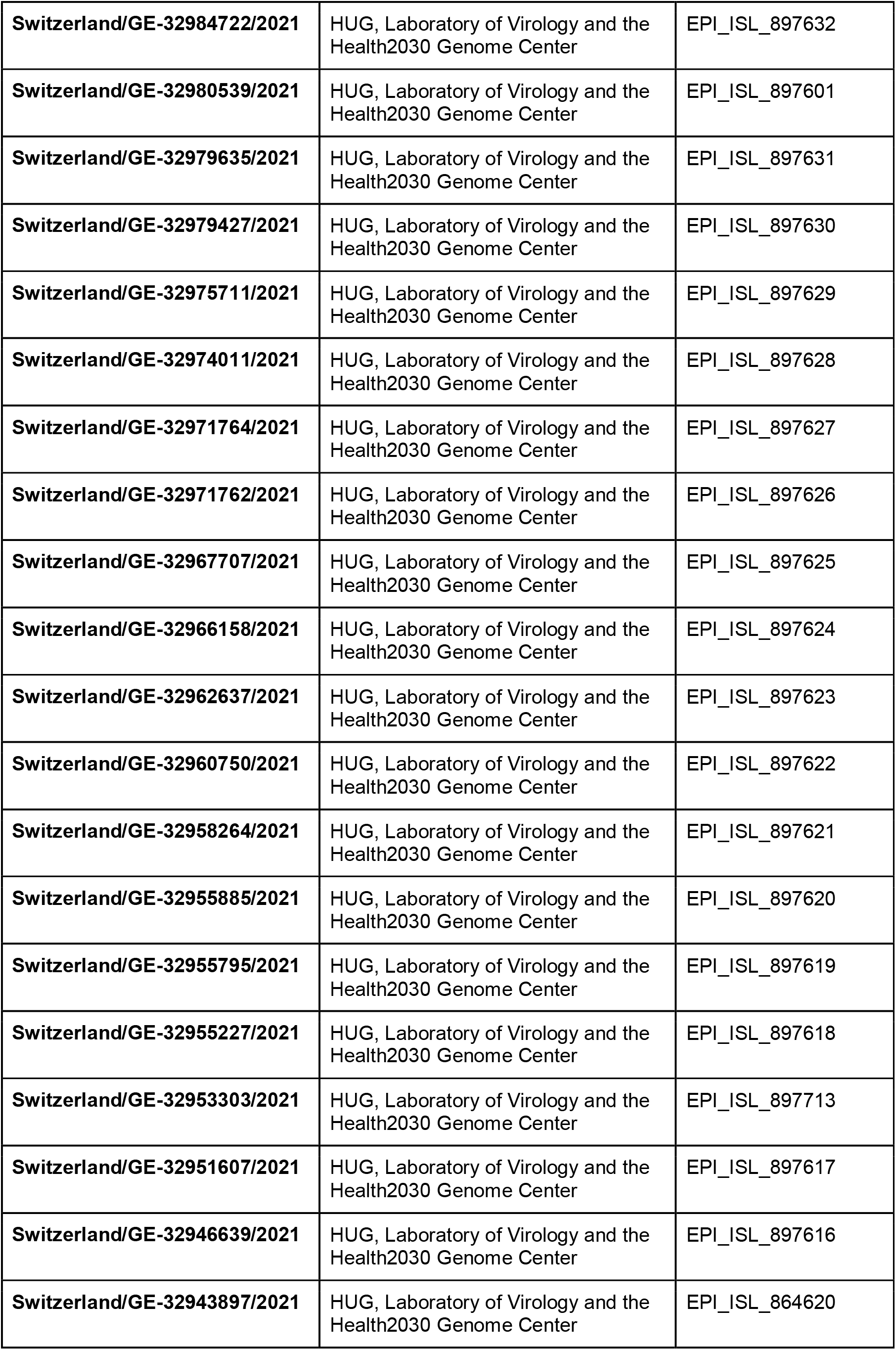
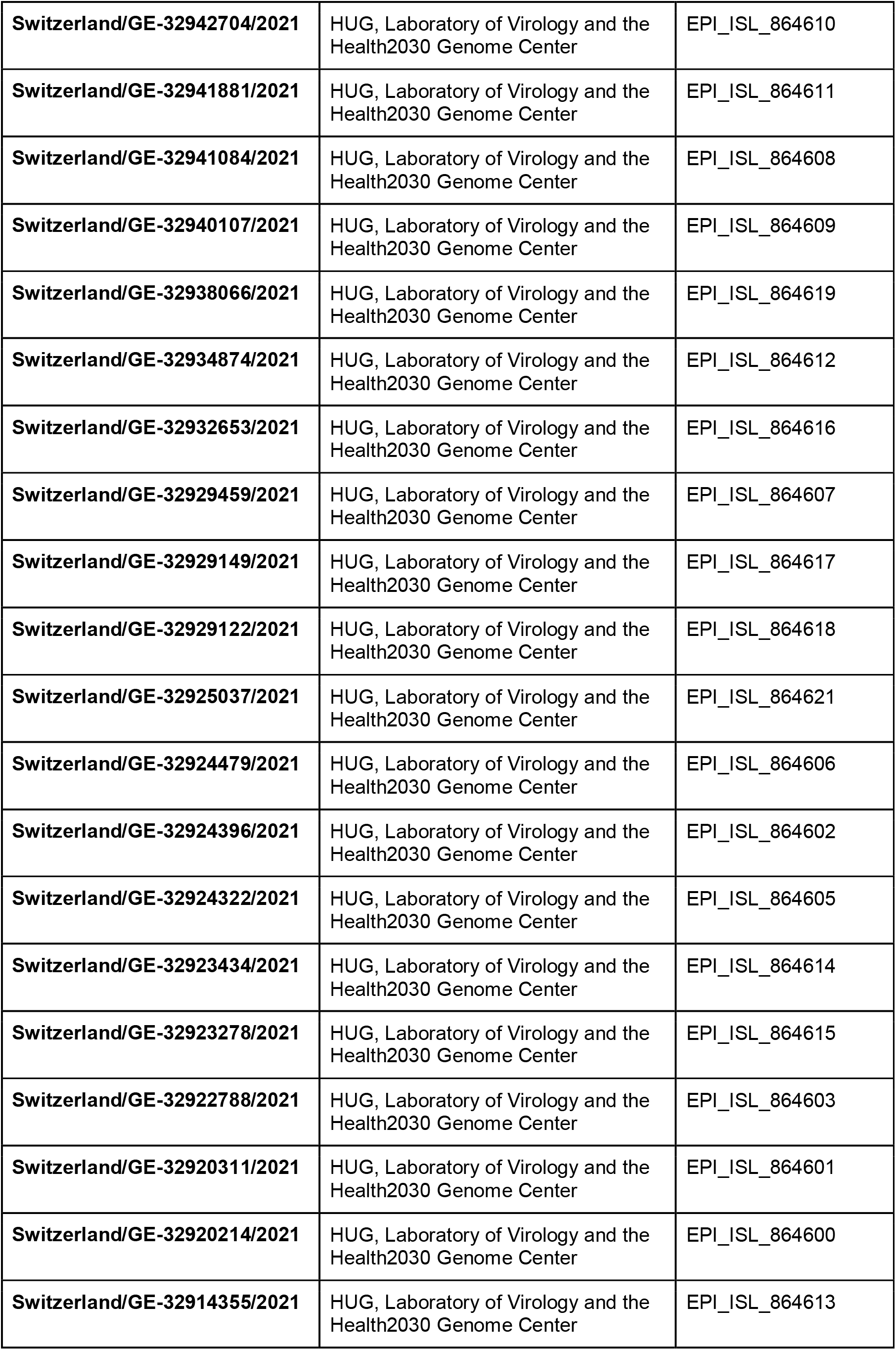
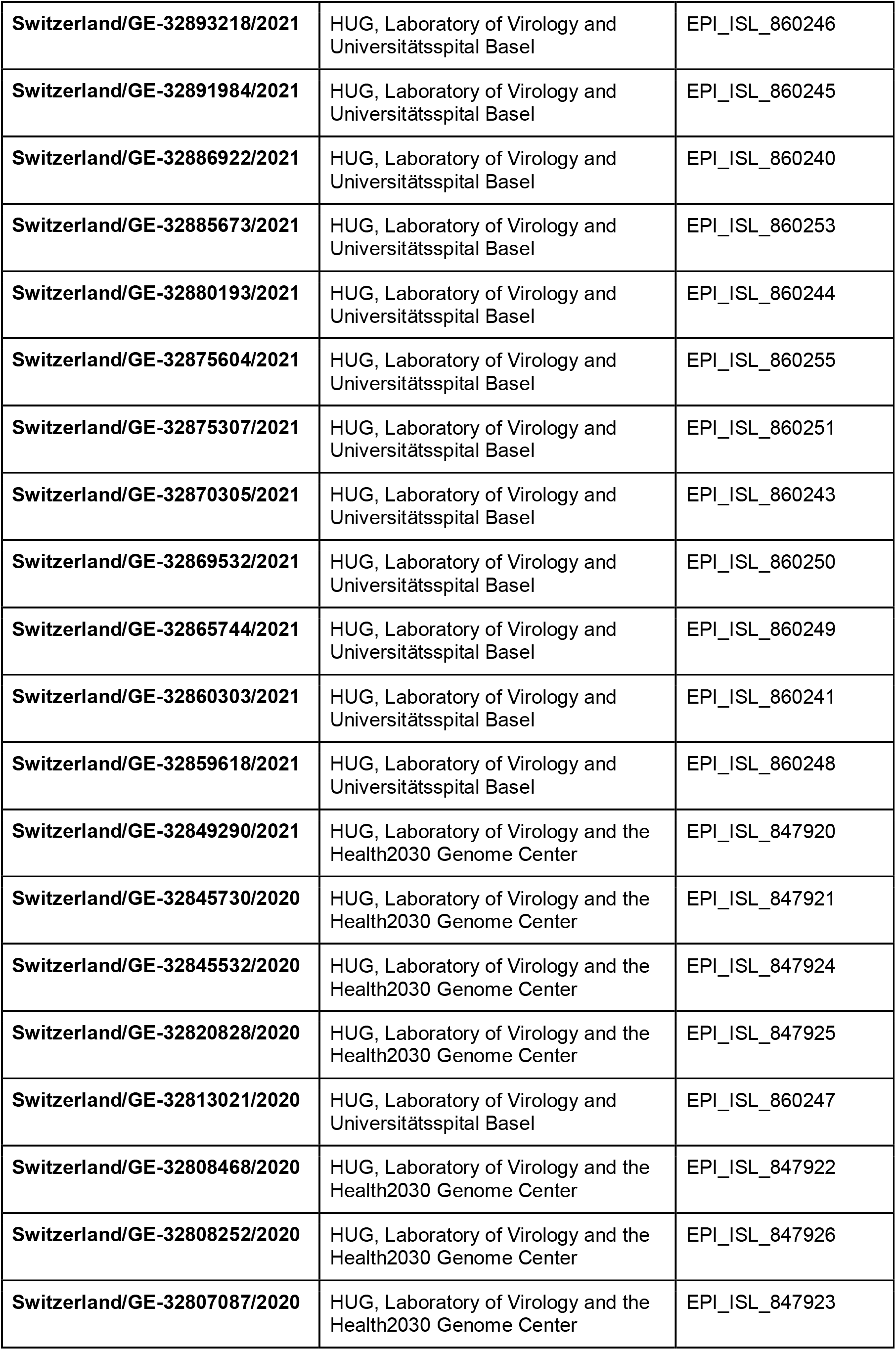
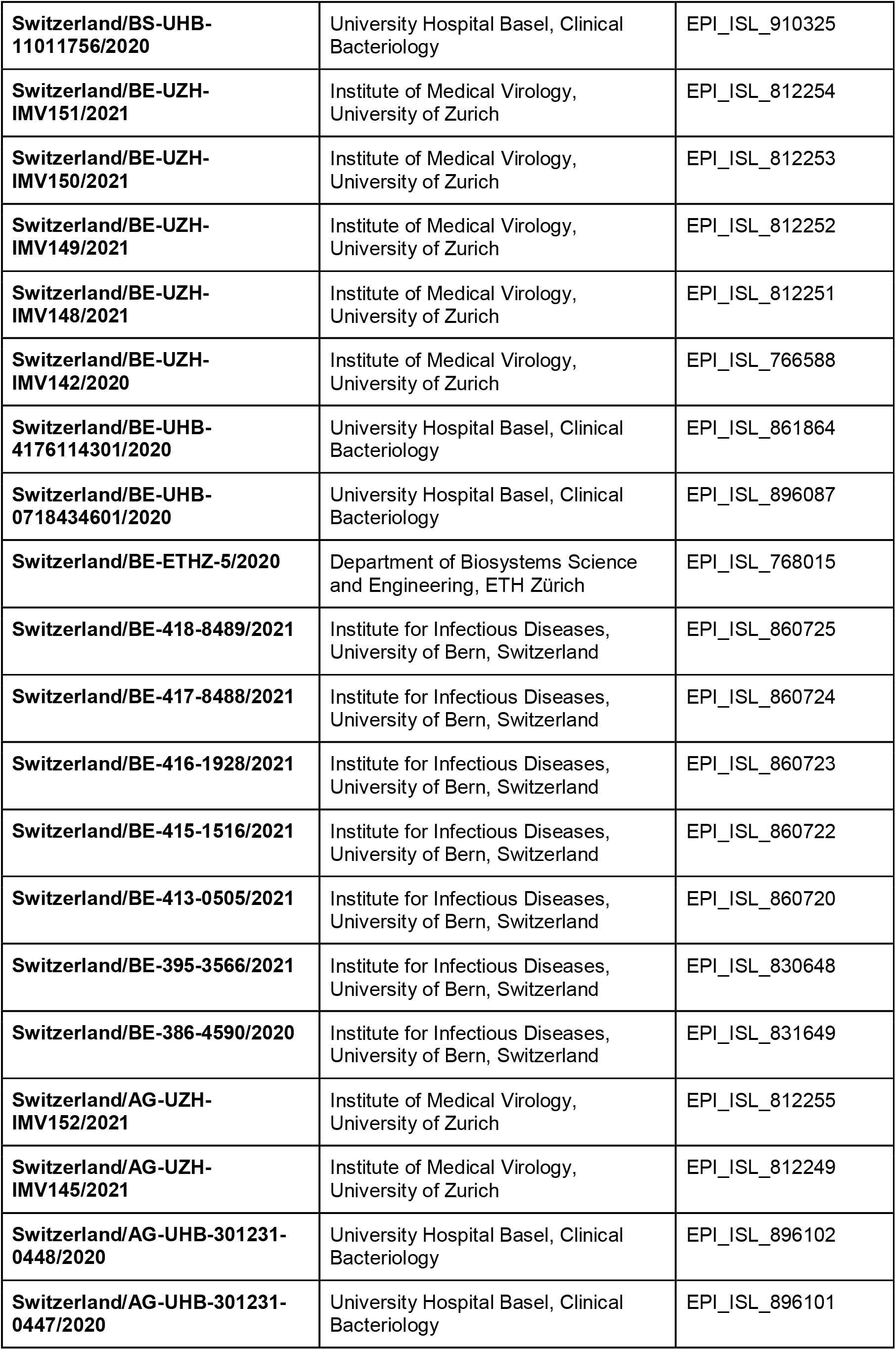
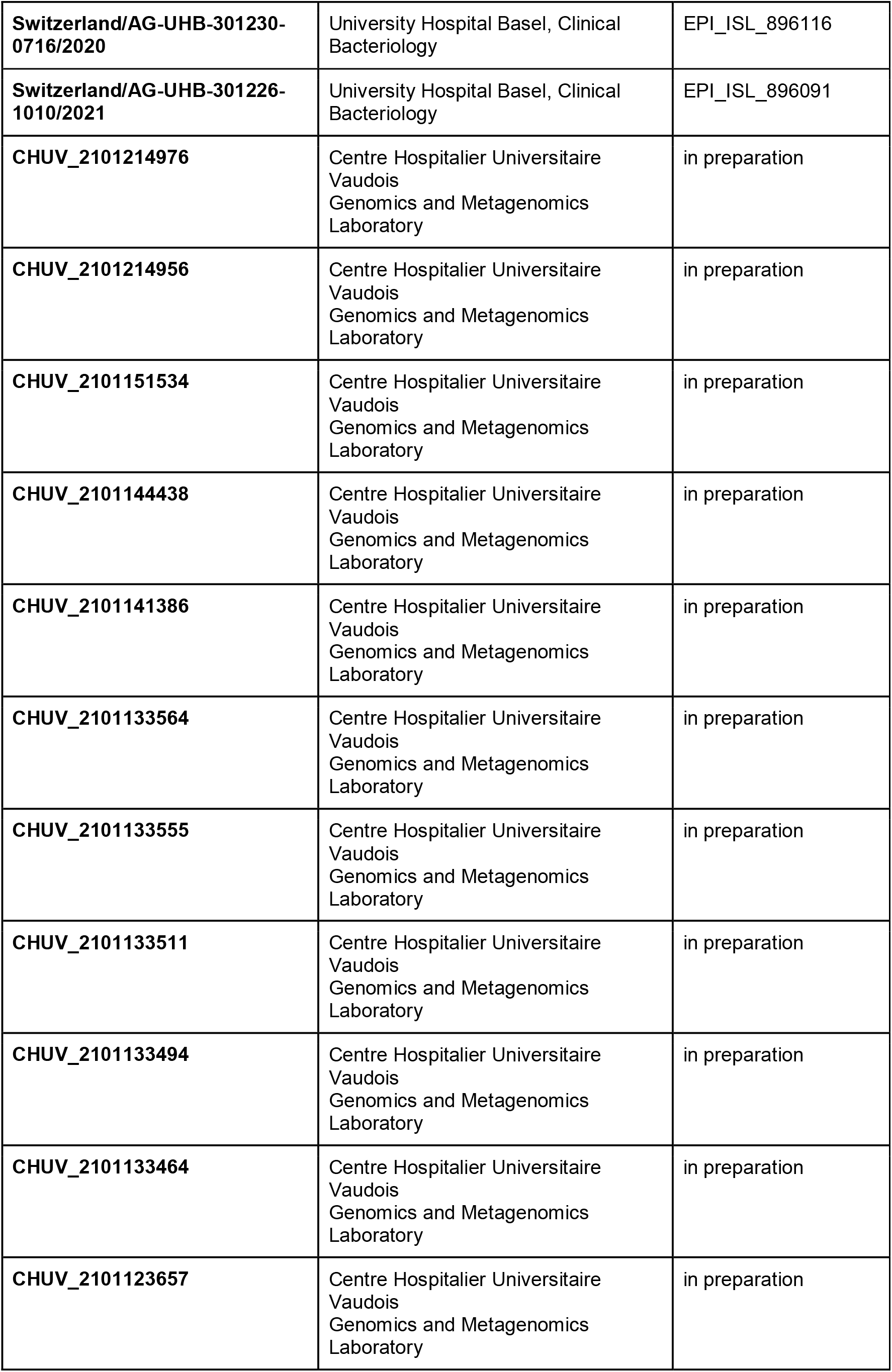
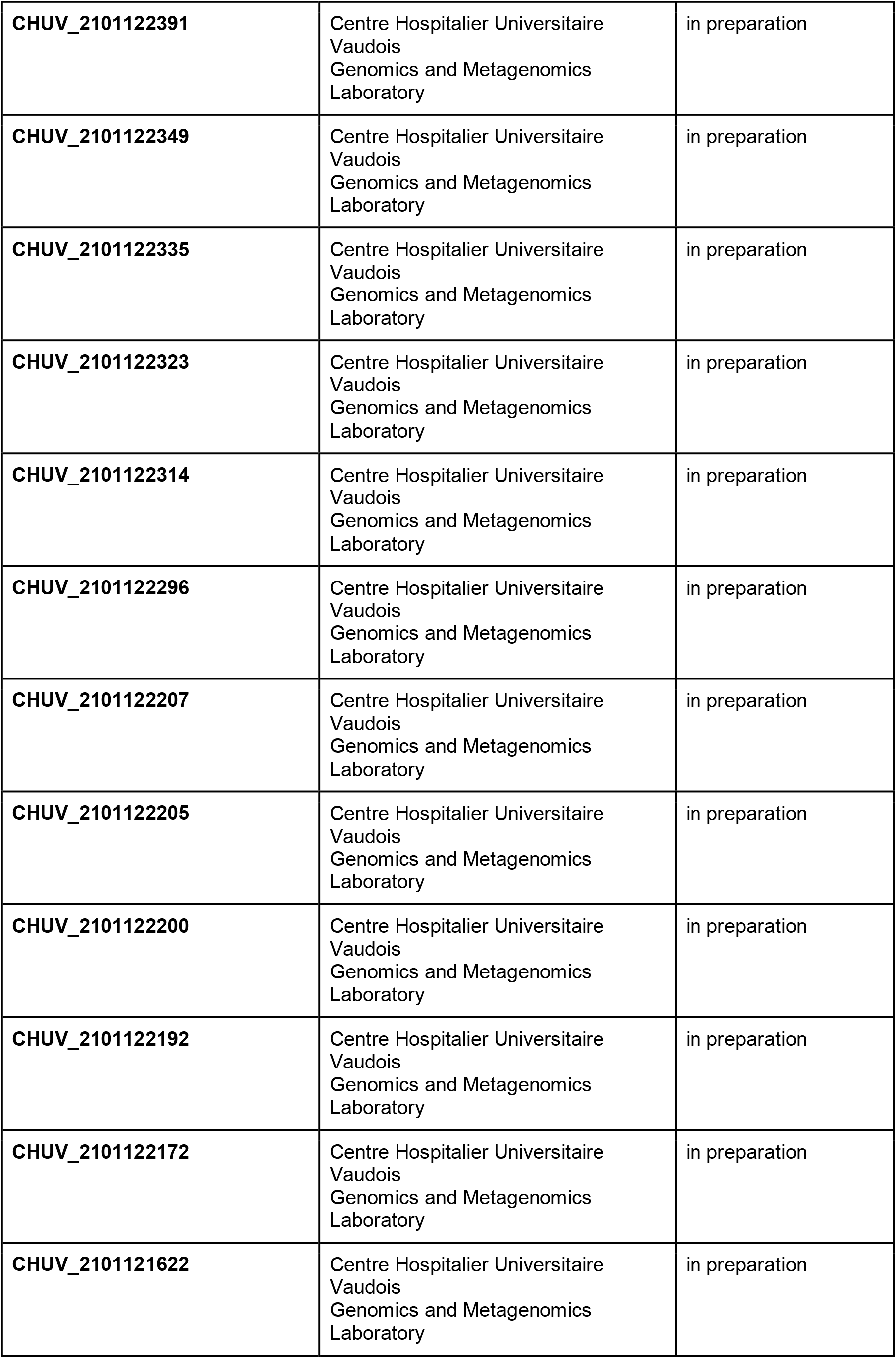
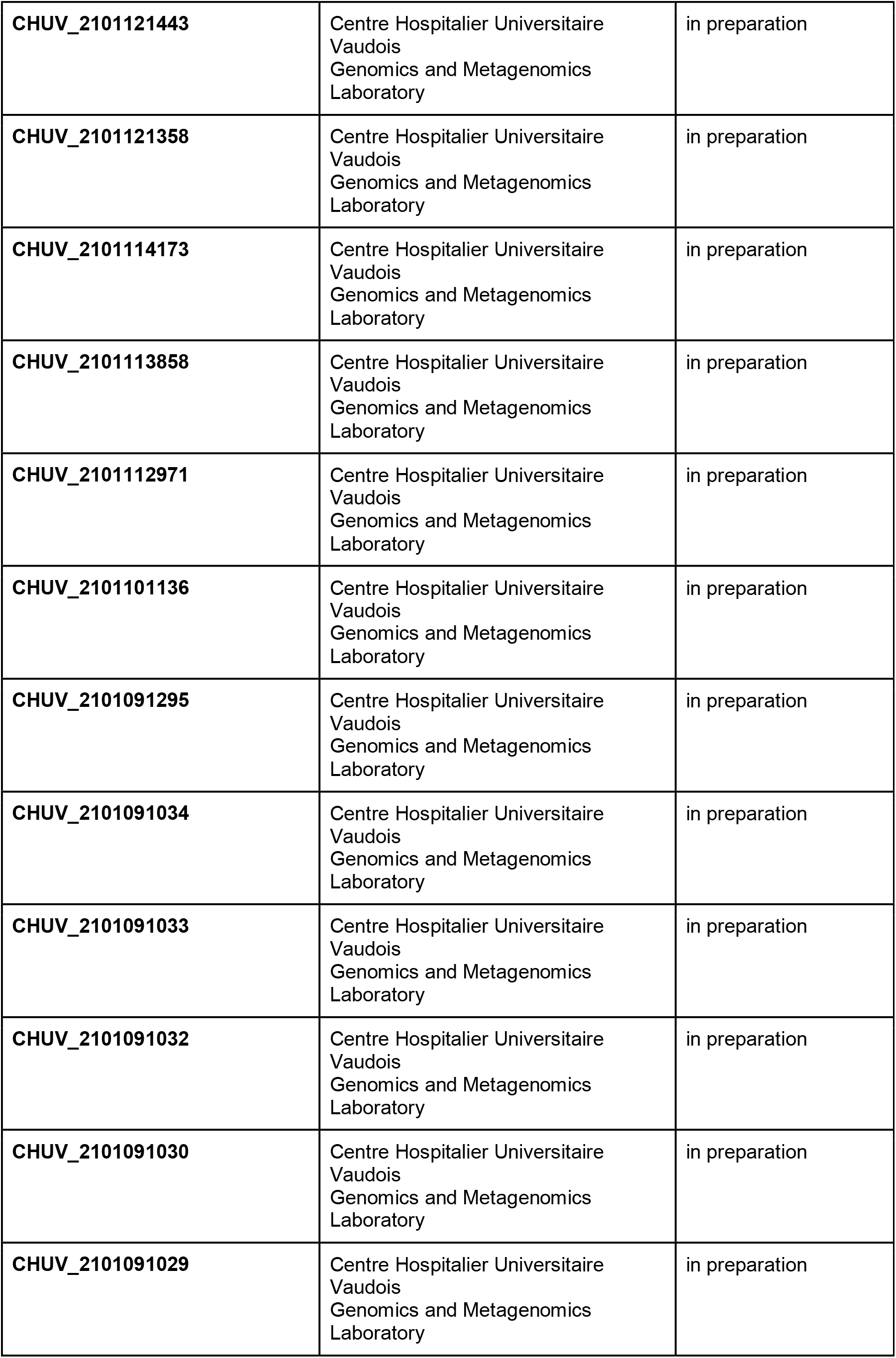
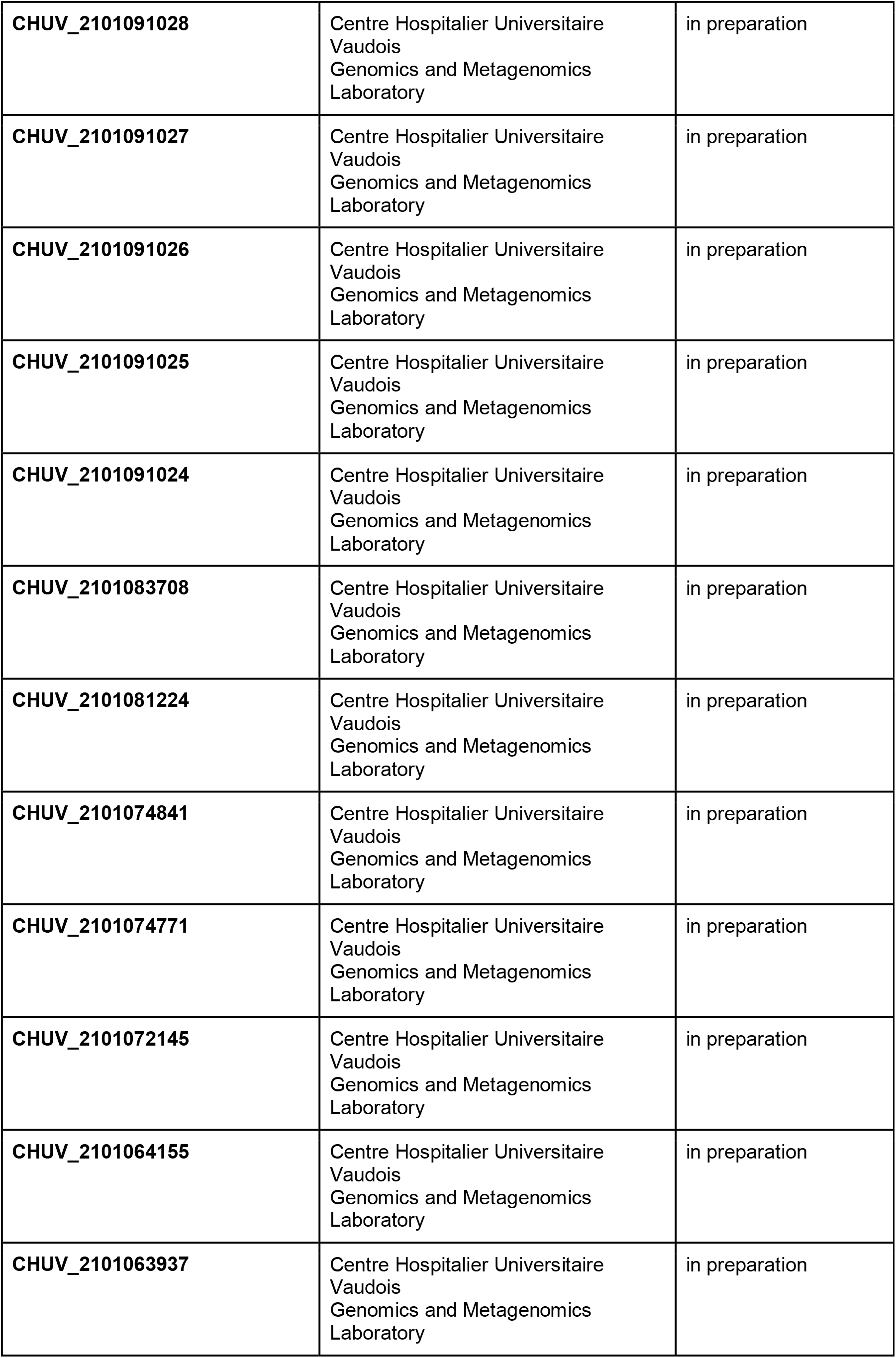
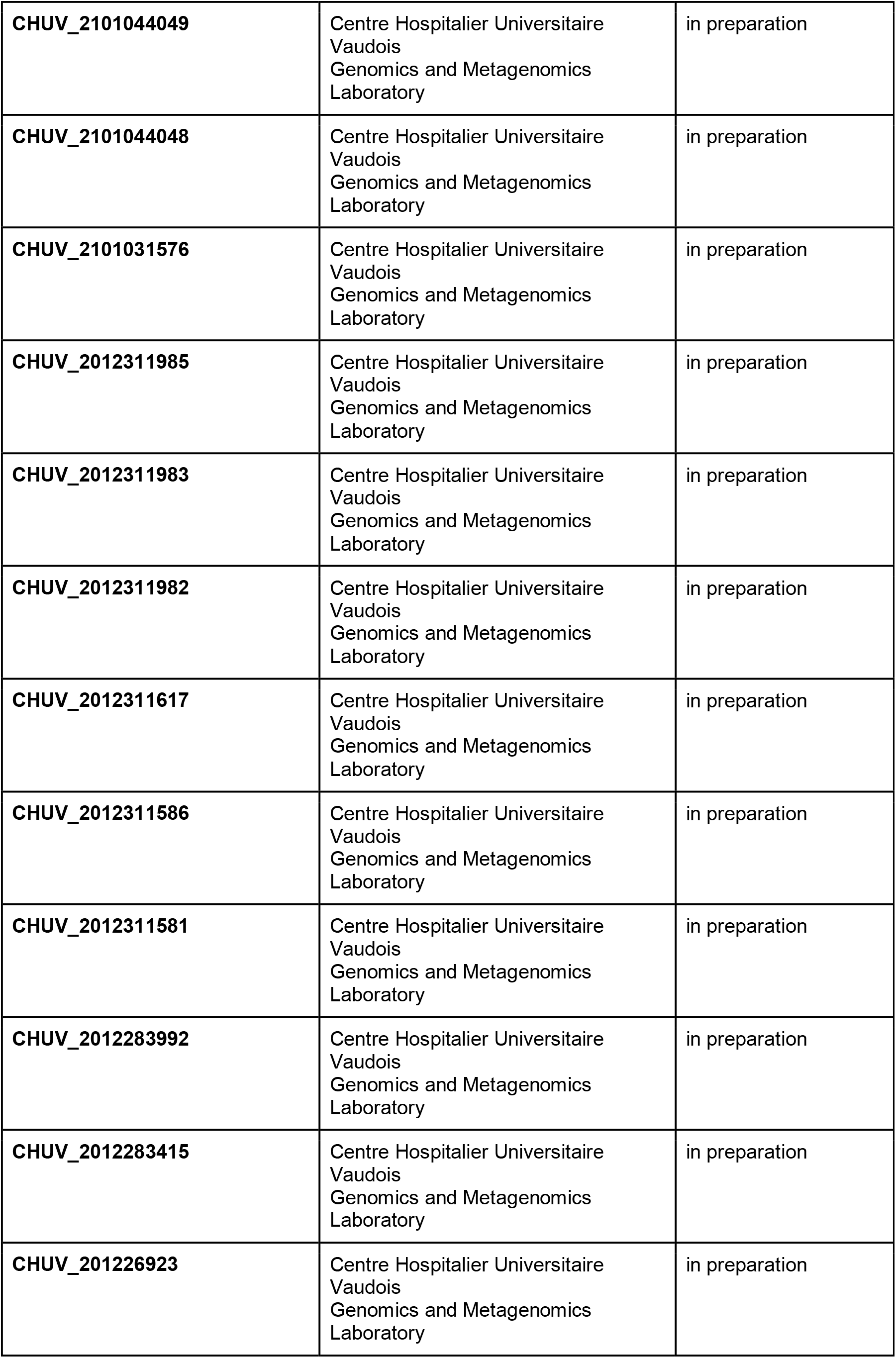
GISAID database identifier. The N501Y-carrying genomes from Switzerland included into phylogenetic analysis are listed.

**Table S5.List of laboratories submitting to GISAID.**We thank all the laboratories actively sharing their datasets.

**Figure S3.**
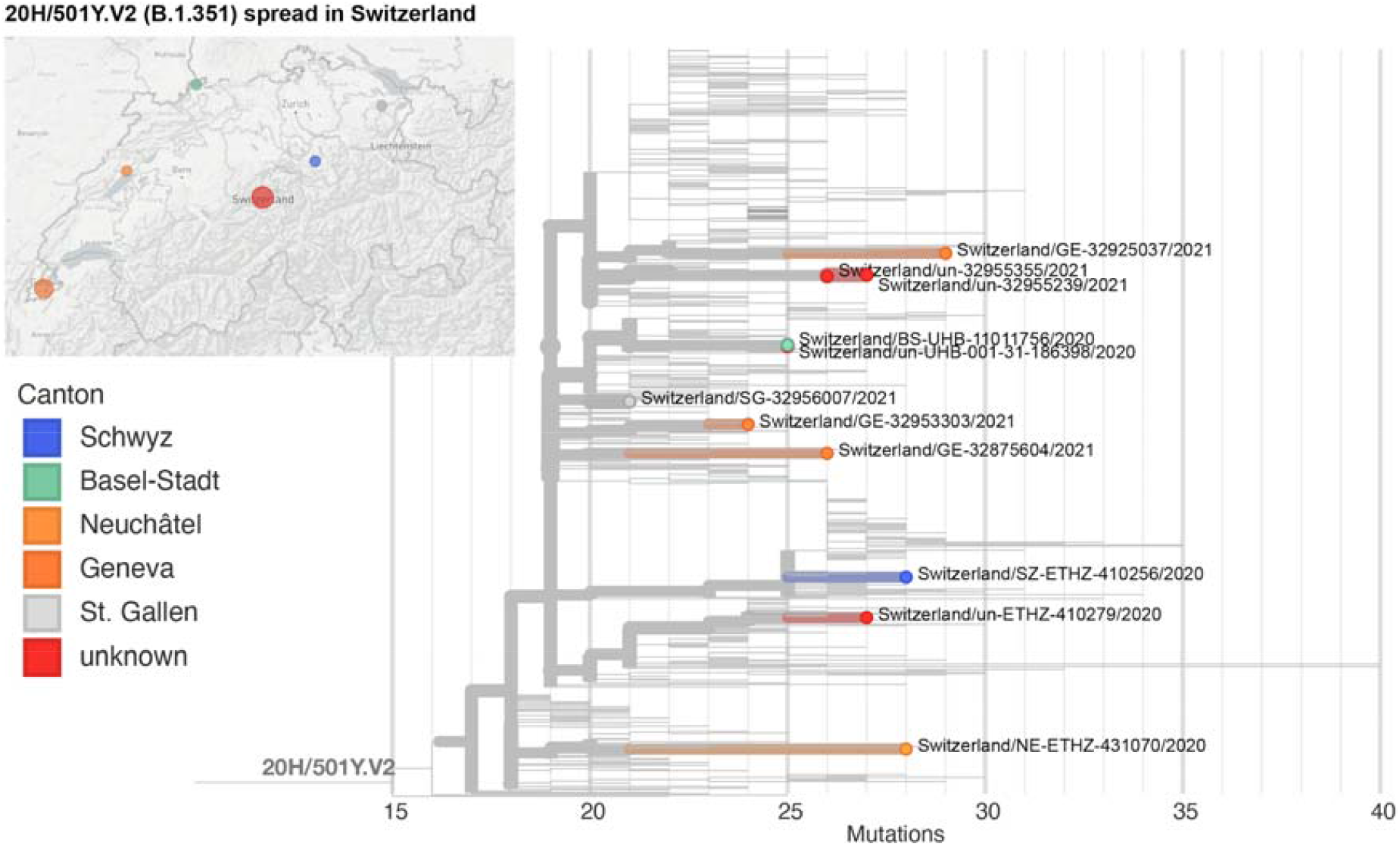
Phylogeny of sequenced B.1.351 cases in Switzerland.

## Acknowledgements

We thank the thank the Federal Office of Public Health for providing the overview data on N501Y-specific PCR results. We thank all diagnostic centers performing the N501Y-specific PCR (as of 02.02.2021) with Laboratoire de virologie (CRIVE-HUG), Institut für medizinische Virologie (University of Zurich), Institut de Microbiologie (University Hospital Lausanne), Institut für Infektionskrankheiten (University of Bern), Klinische Virologie (University Hospital Basel), Analytica Medizinische Laboratorien AG, Bioanalytica AG, Biolytix AG, EOLAB (Bellinzona), Labor Kantonsspital Winterthur, Labor team w AG, Labormedizin SRO AG Spital Langenthal, LMZ Dr. Risch, MCL Medizinisch Laboratorien AG, Medics Labor AG, Synlab (Bioggio and Lausanne), Unilabs (Coppet and Dübendorf), Viollier AG, and Zentrum für Labormedizin (St. Gallen). We thank the sequencing centers for excellent technical assistance: University Hospital Basel with Nadine Blind, Christine Kiessling, Magdalena Schneider, Elisabeth Schultheiss, Clarisse Straub, Daniel Gander, and Rosa-Maria Vesco; University of Bern with Miguel A Terrazos Miani, Stefan Neuenschwander, Cora Sägesser, and Peter Keller; University Hospital Lausanne: Sébastien Aeby, René Brouillet, and Damien Jacot; University of Zurich with Stefan Schmutz, Verena Kufner, Maryam Zaheri, Kevin Steiner, Cyril Shah, Jon Huder, and Jürg Böni. Computations were performed at sciCORE (http://scicore.unibas.ch) scientific computing facility at the University of Basel. We also thank all colleagues submitting data to GISAID - a list of all people contributing used sequences to GISAID is available as **supplementary table 5**.

## Author contributions

Draft of first version: AE, KKS, HSS, MS; PCR and epidemiological data: DP, TS, PB, HHH, KL, OO, GG, MH, AT, FS, MB; samples: CB, LR, NW, GML, LB, IS; sequencing data CC, TS, TR, MS, AE, ARGC, SC, MH, AT, AR, CB, GG; concept development: AE, LK, AT, SL, HHH, GG, TS, MB, JS, CM; bioinformatic tools used: RN, EH, AM, MS, HSS, TR, TP, AL; reviewing of manuscript: all.

