## Supplementary material for "SARS-CoV-2 N501Y introductions and transmissions in Switzerland from beginning of October 2020 to February 2021 – implementation of Swiss-wide diagnostic screening and whole genome sequencing": Table 1

| **Canton** | **B.1.1.7 (501Y.V1)** | **B.1.351 (501Y.V2)** | **Lineage not specified** | **VoC total** |
| --- | --- | --- | --- | --- |
| AG | 40 | 1 | 123 | 164 |
| AI | 1 |  |  | 1 |
| AR |  | 1 | 21 | 22 |
| BE | 232 | 4 | 130 | 366 |
| BL | 64 | 1 | 31 | 96 |
| BS | 29 | 2 | 49 | 80 |
| FR | 69 | 2 | 94 | 165 |
| FL | 28 | 1 | 2 | 31 |
| *GE* | *149* | *4* | *406* | *559* |
| GL | 4 | 1 | 2 | 7 |
| GR | 82 |  | 160 | 242 |
| JU | 105 | 1 | 3 | 109 |
| LU | 12 |  | 86 | 98 |
| NE | 24 |  | 12 | 36 |
| NW | 1 |  | 3 | 4 |
| OW |  |  |  | 0 |
| SG | 52 | 22 | 152 | 226 |
| SH | 9 | 1 | 1 | 11 |
| SO | 43 | 1 | 18 | 62 |
| SZ | 11 | 2 | 27 | 40 |
| TG | 27 | 4 | 36 | 67 |
| TI | 32 | 2 | 133 | 167 |
| UR |  |  |  | 0 |
| VD | 217 | 3 | 48 | 268 |
| VS | 49 |  | 93 | 142 |
| ZG | 2 | 1 | 38 | 41 |
| ZH | 88 | 7 | 393 | 488 |
| **CH/FL** | **1370** | **61** | **2061** | **3492** |

**Table 1.** **Absolute numbers of variants of concern (VoC) including cases of B.1.1.7 and B.1.351 in Switzerland and Principality of Liechtenstein.** No case of P.1 was detected to 05.02.2021. Absolute numbers reflect a biased sample set due to the initial case definitions and biased distribution of diagnostic capacities. The numbers and distributions of lineages is likely biased due to delay in processing and different sequencing capacities.
