## Supplementary material for "SARS-CoV-2 N501Y introductions and transmissions in Switzerland from beginning of October 2020 to February 2021 – implementation of Swiss-wide diagnostic screening and whole genome sequencing": Table S1

| **Description of mutation**  (gene, mutations: effects) | **Position in Genome** | **B.1.1.7**  N501Y.V1 | **B.1.351**  N501Y.V2 | **P.1**  N501Y.V3 |
| --- | --- | --- | --- | --- |
| Spike, T20N: Unknown effects | C21621A | not lineage defining | not lineage defining | **lineage defining** |
| Spike, P26S: Unknown effects | C21638T | not lineage defining | not lineage defining | **lineage defining** |
| Spike, D138Y: Unknown effects | G21974T | not lineage defining | not lineage defining | **lineage defining** |
| Spike, R190S: Unknown effects | G22132T | not lineage defining | not lineage defining | **lineage defining** |
| Spike, N501Y: May bind more tightly to the human angiotensin-converting enzyme 2 (ACE2) receptor | A23063T | **lineage defining** | **lineage defining** | **lineage defining** |
| Spike, H655Y: Unknown effects | C23525T | **not lineage defining** | **not lineage defining** | **lineage defining** |
| Spike, T1027I: Unknown effects | C24642T | **not lineage defining** | **not lineage defining** | **lineage defining** |
| Spike, double deletion (HV 69, 70): Enhances viral infectivity by two-fold, may lead to reduced neutralizing activity of antibodies raised against previous variants | 21765-21770 deletion | **lineage defining** | not lineage defining | not lineage defining |
| Spike, deletion Y144: Confers resistance to 4A8 monoclonal antibody | 21991-21993 deletion | **lineage defining** | not lineage defining | not lineage defining |
| Spike, P681H: Adjacent to the furin cleavage site, may plausibly affect transmissibility | C23604A | **lineage defining** | not lineage defining | not lineage defining |
| Spike, D614G: Already dominant world-wide | A23403G | **lineage defining** | **lineage defining** | not lineage defining |
| Spike, E484K: Leads to reduced neutralizing activity of antibodies raised against previous variants, may increase affinity for ACEII | G23012A | not lineage defining | **lineage defining** | **lineage defining** |
| Spike, A570D, T716I, S982A, D1118H: Unknown effects | C23271A, C23709T, T24506G, G24914C | **lineage defining** | **not lineage defining** | **not lineage defining** |
| Spike, L18F, K417N: Unknown effects | C21614T, G22813T | **not lineage defining** | **lineage defining** | **lineage defining** |
| Spike, D80A, D215G, R246I, A701V: Unknown effects | A21801C, A22206G, G22299T, C23664T | **not lineage defining** | **lineage defining** | not lineage defining |
| ORF1ab, triple deletion SGF 3675-3677: Unknown effects | 11288-11296 deletion | **lineage defining** | not lineage defining | **lineage defining** |
| ORF1ab, K1795Q: Unknown effects | A5648C | not lineage defining | not lineage defining | **lineage defining** |
| ORF1ab, T1001I, A1708D, I2230T: Unknown effects | C3267T, C5388A, T6954C | **lineage defining** | not lineage defining | not lineage defining |
| ORF1ab, E5665D: Unknown effects | G17259T | not lineage defining | not lineage defining | **lineage defining** |
| ORF8, Q27Stop: Early stop codon likely to render ORF8 non-functional. ORF8 deletions/mutations are associated with milder clinical course and lower post-infection inflammation. ORF8 is involved in immune evasion by down-regulation of MHC class 1 | C27972T | **lineage defining** | not lineage defining | not lineage defining |
| ORF8, R52I, Y73C: Likely irrelevant due to earlier stop codon | G28048T, A28111G | **lineage defining** | not lineage defining | not lineage defining |
| ORF7a, E92K: Unknown effects | G27667A | not lineage defining | not lineage defining | **lineage defining** |
| ORF8, insertion 28269-28273: Unknown effects | ins28269-28273 | not lineage defining | not lineage defining | **lineage defining** |
| N, P80R: Unknown effects | C28512G | not lineage defining | not lineage defining | **lineage defining** |
| N, D3L, S235F: Unknown effects | 28280 GAT->CTA, C28977T | **lineage defining** | not lineage defining | not lineage defining |

**Table S1.** Mutations of the new SARS-CoV-2 lineages B.1.1.7, B.1.351, and P.1. Data from GISAID and Covariants.org/shared-mutations. Full SARS-CoV-2 genome of Wuhan-Hu-1 is available (https://www.ncbi.nlm.nih.gov/nuccore/MN908947).
