## Supplementary material for "SARS-CoV-2 N501Y introductions and transmissions in Switzerland from beginning of October 2020 to February 2021 – implementation of Swiss-wide diagnostic screening and whole genome sequencing": Table S2

| **Lineage** | **Frequency** | **Countries** | **Number genomes** | **Percentage of**  **N501Y** |
| --- | --- | --- | --- | --- |
| **B.1** | 6 | USA, United Kingdom | 24057 | 0.025 |
| **B.1.1** | 3 | United Kingdom, USA | 21672 | 0.014 |
| **B.1.1.189** | 1 | Denmark | 113 | 0.885 |
| **B.1.1.7** | 3072 | Australia, Canada, Denmark, United Kingdom, Finland, Hong Kong, India, Ireland, Israel, Italy, Netherlands, Norway, Portugal, Singapore, Spain | 3155 | 97.369 |
| **B.1.1.70** | 484 | United Kingdom | 1009 | 47.968 |
| **B.1.160** | 1 | United Kingdom | 4536 | 0.022 |
| **B.1.177** | 6 | United Kingdom | 35019 | 0.017 |
| **B.1.351** | 326 | United Kingdom, South Africa, Switzerland | 329 | 99.088 |
| **B.1.5** | 9 | United Kingdom | 10533 | 0.085 |
| **B.1.83** | 1 | United Kingdom | 25 | 4 |

**Table S2. N501Y mutations across different viral lineages since September 2020.** Based on GISAID database (access 15.01.2021).
