## Supplementary material for "SARS-CoV-2 N501Y introductions and transmissions in Switzerland from beginning of October 2020 to February 2021 – implementation of Swiss-wide diagnostic screening and whole genome sequencing": Table S3

| **Center** | **Initial case definition** | **PCR approach** | **Sequencing method** |
| --- | --- | --- | --- |
| Microbiological Laboratory EOC Bellinzona | Epidemiological | N501Y | Sent to University center |
| LMZ Dr. Risch AG | Epidemiological  S gene dropout | N501Y | Sent to University center |
| University Hospital Basel | Epidemiological | N501Y  (additional in house method) | Illumina and Oxford Nanopore based whole genome sequencing |
| University of Bern | Epidemiological | N501Y | Oxford Nanopore based amplicon and whole genome sequencing |
| University Hospital Geneva | Epidemiological  S gene dropout | N501Y | Amplicon based Sanger sequencing and WGS |
| University Hospital Lausanne | Epidemiological  S gene dropout | N501Y  (additional in house method, i.e. targeting ORF8) | Illumina based whole genome sequencing |
| University of Zurich | Epidemiological | N501Y | Illumina based whole genome sequencing |
| Centre for Laboratory Medicine St. Gall | Epidemiological | N501Y | Illumina based whole genome sequencing |

**Table S3.** **Screening methods used by different diagnostic laboratories as of 20th January 2021.** N501Y screening assay was most commonly used from TIB MOLBIOL (Germany), but also in-house developed assays were used. University centers offering whole genome sequencing included the University Hospital Basel (Division of Clinical Bacteriology and Mycology), University of Bern (Institute for Infectious Diseases), University Hospital Geneva (Virology Laboratory), University Hospital Lausanne (Institute of Microbiology), and the University of Zurich (Institute of Medical Virology). In addition, ETHZ (D-BSSE core facility) sequenced samples.
