## Supplementary material for "SARS-CoV-2 N501Y introductions and transmissions in Switzerland from beginning of October 2020 to February 2021 – implementation of Swiss-wide diagnostic screening and whole genome sequencing": Table S4

| **GISAID identifier** | **Submitting lab** | **EPI_ISL** |
| --- | --- | --- |
| **Switzerland/ZH-UZH-IMV154/2021** | Institute of Medical Virology, University of Zurich | EPI_ISL_812257 |
| **Switzerland/ZH-UZH-IMV146/2020** | Institute of Medical Virology, University of Zurich | EPI_ISL_812250 |
| **Switzerland/ZH-UZH-IMV130/2020** | Institute of Medical Virology, University of Zurich | EPI_ISL_751193 |
| **Switzerland/ZH-UHB-718502801/2020** | University Hospital Basel, Clinical Bacteriology | EPI_ISL_896104 |
| **Switzerland/ZH-UHB-717847701/2020** | University Hospital Basel, Clinical Bacteriology | EPI_ISL_861862 |
| **Switzerland/ZH-UHB-717559301/2020** | University Hospital Basel, Clinical Bacteriology | EPI_ISL_861861 |
| **Switzerland/ZH-SNRCI-32927909/2021** | HUG, Laboratory of Virology and the Health2030 Genome Center | EPI_ISL_864737 |
| **Switzerland/VS-33014410/2021** | HUG, Laboratory of Virology and the Health2030 Genome Center | EPI_ISL_897679 |
| **Switzerland/VS-33007317/2021** | HUG, Laboratory of Virology and the Health2030 Genome Center | EPI_ISL_897613 |
| **Switzerland/VS-33007285/2021** | HUG, Laboratory of Virology and the Health2030 Genome Center | EPI_ISL_897678 |
| **Switzerland/VS-32987160/2021** | HUG, Laboratory of Virology and the Health2030 Genome Center | EPI_ISL_897677 |
| **Switzerland/VS-32970201/2021** | HUG, Laboratory of Virology and the Health2030 Genome Center | EPI_ISL_897598 |
| **Switzerland/VS-32956055/2021** | HUG, Laboratory of Virology and the Health2030 Genome Center | EPI_ISL_897676 |
| **Switzerland/VS-32867209/2021** | HUG, Laboratory of Virology and Universitätsspital Basel | EPI_ISL_860252 |
| **Switzerland/VS-32859876/2021** | HUG, Laboratory of Virology and Universitätsspital Basel | EPI_ISL_860254 |
| **Switzerland/VD-UZH-IMV144/2020** | Institute of Medical Virology, University of Zurich | EPI_ISL_766590 |
| **Switzerland/VD-UZH-IMV143/2020** | Institute of Medical Virology, University of Zurich | EPI_ISL_766589 |
| **Switzerland/VD-UHB-717443100/2020** | University Hospital Basel, Clinical Bacteriology | EPI_ISL_861859 |
| **Switzerland/VD-UHB-4176481401/2020** | University Hospital Basel, Clinical Bacteriology | EPI_ISL_896088 |
| **Switzerland/VD-UHB-0718440401/2020** | University Hospital Basel, Clinical Bacteriology | EPI_ISL_896103 |
| **Switzerland/VD-SNRCI-32961024/2021** | HUG, Laboratory of Virology and the Health2030 Genome Center | EPI_ISL_897603 |
| **Switzerland/VD-32938885/2021** | HUG, Laboratory of Virology and the Health2030 Genome Center | EPI_ISL_864734 |
| **Switzerland/un-UHB-720584600/2021** | University Hospital Basel, Clinical Bacteriology | EPI_ISL_896099 |
| **Switzerland/un-UHB-720271701/2021** | University Hospital Basel, Clinical Bacteriology | EPI_ISL_896112 |
| **Switzerland/un-UHB-719559501/2021** | University Hospital Basel, Clinical Bacteriology | EPI_ISL_896089 |
| **Switzerland/un-UHB-716224001/2020** | University Hospital Basel, Clinical Bacteriology | EPI_ISL_861858 |
| **Switzerland/un-UHB-716122601/2020** | University Hospital Basel, Clinical Bacteriology | EPI_ISL_861857 |
| **Switzerland/un-UHB-4177791901/2021** | University Hospital Basel, Clinical Bacteriology | EPI_ISL_896069 |
| **Switzerland/un-UHB-4177635701/2021** | University Hospital Basel, Clinical Bacteriology | EPI_ISL_896090 |
| **Switzerland/un-UHB-4177450201/2021** | University Hospital Basel, Clinical Bacteriology | EPI_ISL_896084 |
| **Switzerland/un-UHB-4176940001/2021** | University Hospital Basel, Clinical Bacteriology | EPI_ISL_896077 |
| **Switzerland/un-UHB-4176935301/2021** | University Hospital Basel, Clinical Bacteriology | EPI_ISL_896083 |
| **Switzerland/un-UHB-4176923001/2021** | University Hospital Basel, Clinical Bacteriology | EPI_ISL_896097 |
| **Switzerland/un-UHB-4176911401/2021** | University Hospital Basel, Clinical Bacteriology | EPI_ISL_896082 |
| **Switzerland/un-UHB-4176908401/2021** | University Hospital Basel, Clinical Bacteriology | EPI_ISL_896081 |
| **Switzerland/un-UHB-4176713101/2021** | University Hospital Basel, Clinical Bacteriology | EPI_ISL_896073 |
| **Switzerland/un-UHB-4175179401/2020** | University Hospital Basel, Clinical Bacteriology | EPI_ISL_861856 |
| **Switzerland/un-UHB-31187644/2020** | University Hospital Basel, Clinical Bacteriology | EPI_ISL_910030 |
| **Switzerland/un-UHB-31177838/2020** | University Hospital Basel, Clinical Bacteriology | EPI_ISL_910324 |
| **Switzerland/un-UHB-31177835/2020** | University Hospital Basel, Clinical Bacteriology | EPI_ISL_910222 |
| **Switzerland/un-UHB-31028197/2020** | University Hospital Basel, Clinical Bacteriology | EPI_ISL_896094 |
| **Switzerland/un-UHB-30846237/2020** | University Hospital Basel, Clinical Bacteriology | EPI_ISL_896093 |
| **Switzerland/un-UHB-30830994/2020** | University Hospital Basel, Clinical Bacteriology | EPI_ISL_896107 |
| **Switzerland/un-UHB-11020688/2021** | University Hospital Basel, Clinical Bacteriology | EPI_ISL_896070 |
| **Switzerland/un-UHB-11016146/2021** | University Hospital Basel, Clinical Bacteriology | EPI_ISL_896098 |
| **Switzerland/un-UHB-100409625701/2020** | University Hospital Basel, Clinical Bacteriology | EPI_ISL_896095 |
| **Switzerland/un-UHB-0719562301/2021** | University Hospital Basel, Clinical Bacteriology | EPI_ISL_896071 |
| **Switzerland/un-UHB-0719336901/2021** | University Hospital Basel, Clinical Bacteriology | EPI_ISL_896106 |
| **Switzerland/un-UHB-0719165700/2021** | University Hospital Basel, Clinical Bacteriology | EPI_ISL_896080 |
| **Switzerland/un-UHB-0719087200/2021** | University Hospital Basel, Clinical Bacteriology | EPI_ISL_896079 |
| **Switzerland/un-UHB-0718975401/2021** | University Hospital Basel, Clinical Bacteriology | EPI_ISL_896096 |
| **Switzerland/un-UHB-016-31-214802/2020** | University Hospital Basel, Clinical Bacteriology | EPI_ISL_896072 |
| **Switzerland/un-UHB-001-312-23781/2021** | University Hospital Basel, Clinical Bacteriology | EPI_ISL_896075 |
| **Switzerland/un-UHB-001-312-23775/2021** | University Hospital Basel, Clinical Bacteriology | EPI_ISL_896110 |
| **Switzerland/un-UHB-001-31-320558/2021** | University Hospital Basel, Clinical Bacteriology | EPI_ISL_896100 |
| **Switzerland/un-UHB-001-31-319867/2021** | University Hospital Basel, Clinical Bacteriology | EPI_ISL_896111 |
| **Switzerland/un-UHB-001-31-315355/2021** | University Hospital Basel, Clinical Bacteriology | EPI_ISL_896086 |
| **Switzerland/un-UHB-001-31-302300/2021** | University Hospital Basel, Clinical Bacteriology | EPI_ISL_896085 |
| **Switzerland/un-UHB-001-31-274403/2021** | University Hospital Basel, Clinical Bacteriology | EPI_ISL_896113 |
| **Switzerland/un-UHB-001-31-237034/2021** | University Hospital Basel, Clinical Bacteriology | EPI_ISL_896078 |
| **Switzerland/un-UHB-001-31-224049/2021** | University Hospital Basel, Clinical Bacteriology | EPI_ISL_896076 |
| **Switzerland/un-UHB-001-31-186398/2020** | University Hospital Basel, Clinical Bacteriology | EPI_ISL_896114 |
| **Switzerland/un-ETHZ-410279/2020** | Department of Biosystems Science and Engineering, ETH Zürich | EPI_ISL_737642 |
| **Switzerland/un-ETHZ-2/2020** | Department of Biosystems Science and Engineering, ETH Zürich | EPI_ISL_811130 |
| **Switzerland/un-33001761/2021** | HUG, Laboratory of Virology and the Health2030 Genome Center | EPI_ISL_897675 |
| **Switzerland/un-32987164/2021** | HUG, Laboratory of Virology and the Health2030 Genome Center | EPI_ISL_897674 |
| **Switzerland/un-32986844/2021** | HUG, Laboratory of Virology and the Health2030 Genome Center | EPI_ISL_897673 |
| **Switzerland/un-32955355/2021** | HUG, Laboratory of Virology and the Health2030 Genome Center | EPI_ISL_897604 |
| **Switzerland/un-32955239/2021** | HUG, Laboratory of Virology and the Health2030 Genome Center | EPI_ISL_897714 |
| **Switzerland/TI-33010784/2021** | HUG, Laboratory of Virology and the Health2030 Genome Center | EPI_ISL_897672 |
| **Switzerland/TI-33010582/2021** | HUG, Laboratory of Virology and the Health2030 Genome Center | EPI_ISL_897671 |
| **Switzerland/TI-33009173/2021** | HUG, Laboratory of Virology and the Health2030 Genome Center | EPI_ISL_897670 |
| **Switzerland/SZ-UHB-100409126701/2020** | University Hospital Basel, Clinical Bacteriology | EPI_ISL_861866 |
| **Switzerland/SZ-UHB-100408915101/2020** | University Hospital Basel, Clinical Bacteriology | EPI_ISL_861865 |
| **Switzerland/SZ-ETHZ-410256/2020** | Department of Biosystems Science and Engineering, ETH Zürich | EPI_ISL_737488 |
| **Switzerland/SG-UHB-4175525901/2020** | University Hospital Basel, Clinical Bacteriology | EPI_ISL_861863 |
| **Switzerland/SG-32956007/2021** | HUG, Laboratory of Virology and the Health2030 Genome Center | EPI_ISL_897605 |
| **Switzerland/SG-32938747/2021** | HUG, Laboratory of Virology and the Health2030 Genome Center | EPI_ISL_864718 |
| **Switzerland/NE-ETHZ-431070/2020** | Department of Biosystems Science and Engineering, ETH Zürich | EPI_ISL_899175 |
| **Switzerland/GR-UHB-717511401/2020** | University Hospital Basel, Clinical Bacteriology | EPI_ISL_861860 |
| **Switzerland/GE-ETHZ-420394/2020** | Department of Biosystems Science and Engineering, ETH Zürich | EPI_ISL_768105 |
| **Switzerland/GE-33015157/2021** | HUG, Laboratory of Virology and the Health2030 Genome Center | EPI_ISL_897669 |
| **Switzerland/GE-33014209/2021** | HUG, Laboratory of Virology and the Health2030 Genome Center | EPI_ISL_897668 |
| **Switzerland/GE-33012307/2021** | HUG, Laboratory of Virology and the Health2030 Genome Center | EPI_ISL_897667 |
| **Switzerland/GE-33010304/2021** | HUG, Laboratory of Virology and the Health2030 Genome Center | EPI_ISL_897666 |
| **Switzerland/GE-33010190/2021** | HUG, Laboratory of Virology and the Health2030 Genome Center | EPI_ISL_897665 |
| **Switzerland/GE-33009879/2021** | HUG, Laboratory of Virology and the Health2030 Genome Center | EPI_ISL_897664 |
| **Switzerland/GE-33008898/2021** | HUG, Laboratory of Virology and the Health2030 Genome Center | EPI_ISL_897663 |
| **Switzerland/GE-33008755/2021** | HUG, Laboratory of Virology and the Health2030 Genome Center | EPI_ISL_897662 |
| **Switzerland/GE-33008184/2021** | HUG, Laboratory of Virology and the Health2030 Genome Center | EPI_ISL_897661 |
| **Switzerland/GE-33007670/2021** | HUG, Laboratory of Virology and the Health2030 Genome Center | EPI_ISL_897660 |
| **Switzerland/GE-33007562/2021** | HUG, Laboratory of Virology and the Health2030 Genome Center | EPI_ISL_897659 |
| **Switzerland/GE-33002300/2021** | HUG, Laboratory of Virology and the Health2030 Genome Center | EPI_ISL_897658 |
| **Switzerland/GE-33002169/2021** | HUG, Laboratory of Virology and the Health2030 Genome Center | EPI_ISL_897657 |
| **Switzerland/GE-33002031/2021** | HUG, Laboratory of Virology and the Health2030 Genome Center | EPI_ISL_897656 |
| **Switzerland/GE-33000950/2021** | HUG, Laboratory of Virology and the Health2030 Genome Center | EPI_ISL_897655 |
| **Switzerland/GE-33000809/2021** | HUG, Laboratory of Virology and the Health2030 Genome Center | EPI_ISL_897654 |
| **Switzerland/GE-33000775/2021** | HUG, Laboratory of Virology and the Health2030 Genome Center | EPI_ISL_897653 |
| **Switzerland/GE-33000673/2021** | HUG, Laboratory of Virology and the Health2030 Genome Center | EPI_ISL_897652 |
| **Switzerland/GE-32998367/2021** | HUG, Laboratory of Virology and the Health2030 Genome Center | EPI_ISL_897651 |
| **Switzerland/GE-32997830/2021** | HUG, Laboratory of Virology and the Health2030 Genome Center | EPI_ISL_897650 |
| **Switzerland/GE-32997521/2021** | HUG, Laboratory of Virology and the Health2030 Genome Center | EPI_ISL_897649 |
| **Switzerland/GE-32996759/2021** | HUG, Laboratory of Virology and the Health2030 Genome Center | EPI_ISL_897648 |
| **Switzerland/GE-32996014/2021** | HUG, Laboratory of Virology and the Health2030 Genome Center | EPI_ISL_897647 |
| **Switzerland/GE-32990906/2021** | HUG, Laboratory of Virology and the Health2030 Genome Center | EPI_ISL_897615 |
| **Switzerland/GE-32990876/2021** | HUG, Laboratory of Virology and the Health2030 Genome Center | EPI_ISL_897646 |
| **Switzerland/GE-32990811/2021** | HUG, Laboratory of Virology and the Health2030 Genome Center | EPI_ISL_897645 |
| **Switzerland/GE-32990664/2021** | HUG, Laboratory of Virology and the Health2030 Genome Center | EPI_ISL_897614 |
| **Switzerland/GE-32990272/2021** | HUG, Laboratory of Virology and the Health2030 Genome Center | EPI_ISL_897644 |
| **Switzerland/GE-32988968/2021** | HUG, Laboratory of Virology and the Health2030 Genome Center | EPI_ISL_897642 |
| **Switzerland/GE-32987355/2021** | HUG, Laboratory of Virology and the Health2030 Genome Center | EPI_ISL_897641 |
| **Switzerland/GE-32986668/2021** | HUG, Laboratory of Virology and the Health2030 Genome Center | EPI_ISL_897640 |
| **Switzerland/GE-32986564/2021** | HUG, Laboratory of Virology and the Health2030 Genome Center | EPI_ISL_897639 |
| **Switzerland/GE-32986541/2021** | HUG, Laboratory of Virology and the Health2030 Genome Center | EPI_ISL_897638 |
| **Switzerland/GE-32986531/2021** | HUG, Laboratory of Virology and the Health2030 Genome Center | EPI_ISL_897637 |
| **Switzerland/GE-32985530/2021** | HUG, Laboratory of Virology and the Health2030 Genome Center | EPI_ISL_897636 |
| **Switzerland/GE-32985523/2021** | HUG, Laboratory of Virology and the Health2030 Genome Center | EPI_ISL_897635 |
| **Switzerland/GE-32985372/2021** | HUG, Laboratory of Virology and the Health2030 Genome Center | EPI_ISL_897634 |
| **Switzerland/GE-32985291/2021** | HUG, Laboratory of Virology and the Health2030 Genome Center | EPI_ISL_897633 |
| **Switzerland/GE-32984722/2021** | HUG, Laboratory of Virology and the Health2030 Genome Center | EPI_ISL_897632 |
| **Switzerland/GE-32980539/2021** | HUG, Laboratory of Virology and the Health2030 Genome Center | EPI_ISL_897601 |
| **Switzerland/GE-32979635/2021** | HUG, Laboratory of Virology and the Health2030 Genome Center | EPI_ISL_897631 |
| **Switzerland/GE-32979427/2021** | HUG, Laboratory of Virology and the Health2030 Genome Center | EPI_ISL_897630 |
| **Switzerland/GE-32975711/2021** | HUG, Laboratory of Virology and the Health2030 Genome Center | EPI_ISL_897629 |
| **Switzerland/GE-32974011/2021** | HUG, Laboratory of Virology and the Health2030 Genome Center | EPI_ISL_897628 |
| **Switzerland/GE-32971764/2021** | HUG, Laboratory of Virology and the Health2030 Genome Center | EPI_ISL_897627 |
| **Switzerland/GE-32971762/2021** | HUG, Laboratory of Virology and the Health2030 Genome Center | EPI_ISL_897626 |
| **Switzerland/GE-32967707/2021** | HUG, Laboratory of Virology and the Health2030 Genome Center | EPI_ISL_897625 |
| **Switzerland/GE-32966158/2021** | HUG, Laboratory of Virology and the Health2030 Genome Center | EPI_ISL_897624 |
| **Switzerland/GE-32962637/2021** | HUG, Laboratory of Virology and the Health2030 Genome Center | EPI_ISL_897623 |
| **Switzerland/GE-32960750/2021** | HUG, Laboratory of Virology and the Health2030 Genome Center | EPI_ISL_897622 |
| **Switzerland/GE-32958264/2021** | HUG, Laboratory of Virology and the Health2030 Genome Center | EPI_ISL_897621 |
| **Switzerland/GE-32955885/2021** | HUG, Laboratory of Virology and the Health2030 Genome Center | EPI_ISL_897620 |
| **Switzerland/GE-32955795/2021** | HUG, Laboratory of Virology and the Health2030 Genome Center | EPI_ISL_897619 |
| **Switzerland/GE-32955227/2021** | HUG, Laboratory of Virology and the Health2030 Genome Center | EPI_ISL_897618 |
| **Switzerland/GE-32953303/2021** | HUG, Laboratory of Virology and the Health2030 Genome Center | EPI_ISL_897713 |
| **Switzerland/GE-32951607/2021** | HUG, Laboratory of Virology and the Health2030 Genome Center | EPI_ISL_897617 |
| **Switzerland/GE-32946639/2021** | HUG, Laboratory of Virology and the Health2030 Genome Center | EPI_ISL_897616 |
| **Switzerland/GE-32943897/2021** | HUG, Laboratory of Virology and the Health2030 Genome Center | EPI_ISL_864620 |
| **Switzerland/GE-32942704/2021** | HUG, Laboratory of Virology and the Health2030 Genome Center | EPI_ISL_864610 |
| **Switzerland/GE-32941881/2021** | HUG, Laboratory of Virology and the Health2030 Genome Center | EPI_ISL_864611 |
| **Switzerland/GE-32941084/2021** | HUG, Laboratory of Virology and the Health2030 Genome Center | EPI_ISL_864608 |
| **Switzerland/GE-32940107/2021** | HUG, Laboratory of Virology and the Health2030 Genome Center | EPI_ISL_864609 |
| **Switzerland/GE-32938066/2021** | HUG, Laboratory of Virology and the Health2030 Genome Center | EPI_ISL_864619 |
| **Switzerland/GE-32934874/2021** | HUG, Laboratory of Virology and the Health2030 Genome Center | EPI_ISL_864612 |
| **Switzerland/GE-32932653/2021** | HUG, Laboratory of Virology and the Health2030 Genome Center | EPI_ISL_864616 |
| **Switzerland/GE-32929459/2021** | HUG, Laboratory of Virology and the Health2030 Genome Center | EPI_ISL_864607 |
| **Switzerland/GE-32929149/2021** | HUG, Laboratory of Virology and the Health2030 Genome Center | EPI_ISL_864617 |
| **Switzerland/GE-32929122/2021** | HUG, Laboratory of Virology and the Health2030 Genome Center | EPI_ISL_864618 |
| **Switzerland/GE-32925037/2021** | HUG, Laboratory of Virology and the Health2030 Genome Center | EPI_ISL_864621 |
| **Switzerland/GE-32924479/2021** | HUG, Laboratory of Virology and the Health2030 Genome Center | EPI_ISL_864606 |
| **Switzerland/GE-32924396/2021** | HUG, Laboratory of Virology and the Health2030 Genome Center | EPI_ISL_864602 |
| **Switzerland/GE-32924322/2021** | HUG, Laboratory of Virology and the Health2030 Genome Center | EPI_ISL_864605 |
| **Switzerland/GE-32923434/2021** | HUG, Laboratory of Virology and the Health2030 Genome Center | EPI_ISL_864614 |
| **Switzerland/GE-32923278/2021** | HUG, Laboratory of Virology and the Health2030 Genome Center | EPI_ISL_864615 |
| **Switzerland/GE-32922788/2021** | HUG, Laboratory of Virology and the Health2030 Genome Center | EPI_ISL_864603 |
| **Switzerland/GE-32920311/2021** | HUG, Laboratory of Virology and the Health2030 Genome Center | EPI_ISL_864601 |
| **Switzerland/GE-32920214/2021** | HUG, Laboratory of Virology and the Health2030 Genome Center | EPI_ISL_864600 |
| **Switzerland/GE-32914355/2021** | HUG, Laboratory of Virology and the Health2030 Genome Center | EPI_ISL_864613 |
| **Switzerland/GE-32893218/2021** | HUG, Laboratory of Virology and Universitätsspital Basel | EPI_ISL_860246 |
| **Switzerland/GE-32891984/2021** | HUG, Laboratory of Virology and Universitätsspital Basel | EPI_ISL_860245 |
| **Switzerland/GE-32886922/2021** | HUG, Laboratory of Virology and Universitätsspital Basel | EPI_ISL_860240 |
| **Switzerland/GE-32885673/2021** | HUG, Laboratory of Virology and Universitätsspital Basel | EPI_ISL_860253 |
| **Switzerland/GE-32880193/2021** | HUG, Laboratory of Virology and Universitätsspital Basel | EPI_ISL_860244 |
| **Switzerland/GE-32875604/2021** | HUG, Laboratory of Virology and Universitätsspital Basel | EPI_ISL_860255 |
| **Switzerland/GE-32875307/2021** | HUG, Laboratory of Virology and Universitätsspital Basel | EPI_ISL_860251 |
| **Switzerland/GE-32870305/2021** | HUG, Laboratory of Virology and Universitätsspital Basel | EPI_ISL_860243 |
| **Switzerland/GE-32869532/2021** | HUG, Laboratory of Virology and Universitätsspital Basel | EPI_ISL_860250 |
| **Switzerland/GE-32865744/2021** | HUG, Laboratory of Virology and Universitätsspital Basel | EPI_ISL_860249 |
| **Switzerland/GE-32860303/2021** | HUG, Laboratory of Virology and Universitätsspital Basel | EPI_ISL_860241 |
| **Switzerland/GE-32859618/2021** | HUG, Laboratory of Virology and Universitätsspital Basel | EPI_ISL_860248 |
| **Switzerland/GE-32849290/2021** | HUG, Laboratory of Virology and the Health2030 Genome Center | EPI_ISL_847920 |
| **Switzerland/GE-32845730/2020** | HUG, Laboratory of Virology and the Health2030 Genome Center | EPI_ISL_847921 |
| **Switzerland/GE-32845532/2020** | HUG, Laboratory of Virology and the Health2030 Genome Center | EPI_ISL_847924 |
| **Switzerland/GE-32820828/2020** | HUG, Laboratory of Virology and the Health2030 Genome Center | EPI_ISL_847925 |
| **Switzerland/GE-32813021/2020** | HUG, Laboratory of Virology and Universitätsspital Basel | EPI_ISL_860247 |
| **Switzerland/GE-32808468/2020** | HUG, Laboratory of Virology and the Health2030 Genome Center | EPI_ISL_847922 |
| **Switzerland/GE-32808252/2020** | HUG, Laboratory of Virology and the Health2030 Genome Center | EPI_ISL_847926 |
| **Switzerland/GE-32807087/2020** | HUG, Laboratory of Virology and the Health2030 Genome Center | EPI_ISL_847923 |
| **Switzerland/BS-UHB-11011756/2020** | University Hospital Basel, Clinical Bacteriology | EPI_ISL_910325 |
| **Switzerland/BE-UZH-IMV151/2021** | Institute of Medical Virology, University of Zurich | EPI_ISL_812254 |
| **Switzerland/BE-UZH-IMV150/2021** | Institute of Medical Virology, University of Zurich | EPI_ISL_812253 |
| **Switzerland/BE-UZH-IMV149/2021** | Institute of Medical Virology, University of Zurich | EPI_ISL_812252 |
| **Switzerland/BE-UZH-IMV148/2021** | Institute of Medical Virology, University of Zurich | EPI_ISL_812251 |
| **Switzerland/BE-UZH-IMV142/2020** | Institute of Medical Virology, University of Zurich | EPI_ISL_766588 |
| **Switzerland/BE-UHB-4176114301/2020** | University Hospital Basel, Clinical Bacteriology | EPI_ISL_861864 |
| **Switzerland/BE-UHB-0718434601/2020** | University Hospital Basel, Clinical Bacteriology | EPI_ISL_896087 |
| **Switzerland/BE-ETHZ-5/2020** | Department of Biosystems Science and Engineering, ETH Zürich | EPI_ISL_768015 |
| **Switzerland/BE-418-8489/2021** | Institute for Infectious Diseases, University of Bern, Switzerland | EPI_ISL_860725 |
| **Switzerland/BE-417-8488/2021** | Institute for Infectious Diseases, University of Bern, Switzerland | EPI_ISL_860724 |
| **Switzerland/BE-416-1928/2021** | Institute for Infectious Diseases, University of Bern, Switzerland | EPI_ISL_860723 |
| **Switzerland/BE-415-1516/2021** | Institute for Infectious Diseases, University of Bern, Switzerland | EPI_ISL_860722 |
| **Switzerland/BE-413-0505/2021** | Institute for Infectious Diseases, University of Bern, Switzerland | EPI_ISL_860720 |
| **Switzerland/BE-395-3566/2021** | Institute for Infectious Diseases, University of Bern, Switzerland | EPI_ISL_830648 |
| **Switzerland/BE-386-4590/2020** | Institute for Infectious Diseases, University of Bern, Switzerland | EPI_ISL_831649 |
| **Switzerland/AG-UZH-IMV152/2021** | Institute of Medical Virology, University of Zurich | EPI_ISL_812255 |
| **Switzerland/AG-UZH-IMV145/2021** | Institute of Medical Virology, University of Zurich | EPI_ISL_812249 |
| **Switzerland/AG-UHB-301231-0448/2020** | University Hospital Basel, Clinical Bacteriology | EPI_ISL_896102 |
| **Switzerland/AG-UHB-301231-0447/2020** | University Hospital Basel, Clinical Bacteriology | EPI_ISL_896101 |
| **Switzerland/AG-UHB-301230-0716/2020** | University Hospital Basel, Clinical Bacteriology | EPI_ISL_896116 |
| **Switzerland/AG-UHB-301226-1010/2021** | University Hospital Basel, Clinical Bacteriology | EPI_ISL_896091 |
| **CHUV_2101214976** | Centre Hospitalier Universitaire Vaudois  Genomics and Metagenomics Laboratory | in preparation |
| **CHUV_2101214956** | Centre Hospitalier Universitaire Vaudois  Genomics and Metagenomics Laboratory | in preparation |
| **CHUV_2101151534** | Centre Hospitalier Universitaire Vaudois  Genomics and Metagenomics Laboratory | in preparation |
| **CHUV_2101144438** | Centre Hospitalier Universitaire Vaudois  Genomics and Metagenomics Laboratory | in preparation |
| **CHUV_2101141386** | Centre Hospitalier Universitaire Vaudois  Genomics and Metagenomics Laboratory | in preparation |
| **CHUV_2101133564** | Centre Hospitalier Universitaire Vaudois  Genomics and Metagenomics Laboratory | in preparation |
| **CHUV_2101133555** | Centre Hospitalier Universitaire Vaudois  Genomics and Metagenomics Laboratory | in preparation |
| **CHUV_2101133511** | Centre Hospitalier Universitaire Vaudois  Genomics and Metagenomics Laboratory | in preparation |
| **CHUV_2101133494** | Centre Hospitalier Universitaire Vaudois  Genomics and Metagenomics Laboratory | in preparation |
| **CHUV_2101133464** | Centre Hospitalier Universitaire Vaudois  Genomics and Metagenomics Laboratory | in preparation |
| **CHUV_2101123657** | Centre Hospitalier Universitaire Vaudois  Genomics and Metagenomics Laboratory | in preparation |
| **CHUV_2101122391** | Centre Hospitalier Universitaire Vaudois  Genomics and Metagenomics Laboratory | in preparation |
| **CHUV_2101122349** | Centre Hospitalier Universitaire Vaudois  Genomics and Metagenomics Laboratory | in preparation |
| **CHUV_2101122335** | Centre Hospitalier Universitaire Vaudois  Genomics and Metagenomics Laboratory | in preparation |
| **CHUV_2101122323** | Centre Hospitalier Universitaire Vaudois  Genomics and Metagenomics Laboratory | in preparation |
| **CHUV_2101122314** | Centre Hospitalier Universitaire Vaudois  Genomics and Metagenomics Laboratory | in preparation |
| **CHUV_2101122296** | Centre Hospitalier Universitaire Vaudois  Genomics and Metagenomics Laboratory | in preparation |
| **CHUV_2101122207** | Centre Hospitalier Universitaire Vaudois  Genomics and Metagenomics Laboratory | in preparation |
| **CHUV_2101122205** | Centre Hospitalier Universitaire Vaudois  Genomics and Metagenomics Laboratory | in preparation |
| **CHUV_2101122200** | Centre Hospitalier Universitaire Vaudois  Genomics and Metagenomics Laboratory | in preparation |
| **CHUV_2101122192** | Centre Hospitalier Universitaire Vaudois  Genomics and Metagenomics Laboratory | in preparation |
| **CHUV_2101122172** | Centre Hospitalier Universitaire Vaudois  Genomics and Metagenomics Laboratory | in preparation |
| **CHUV_2101121622** | Centre Hospitalier Universitaire Vaudois  Genomics and Metagenomics Laboratory | in preparation |
| **CHUV_2101121443** | Centre Hospitalier Universitaire Vaudois  Genomics and Metagenomics Laboratory | in preparation |
| **CHUV_2101121358** | Centre Hospitalier Universitaire Vaudois  Genomics and Metagenomics Laboratory | in preparation |
| **CHUV_2101114173** | Centre Hospitalier Universitaire Vaudois  Genomics and Metagenomics Laboratory | in preparation |
| **CHUV_2101113858** | Centre Hospitalier Universitaire Vaudois  Genomics and Metagenomics Laboratory | in preparation |
| **CHUV_2101112971** | Centre Hospitalier Universitaire Vaudois  Genomics and Metagenomics Laboratory | in preparation |
| **CHUV_2101101136** | Centre Hospitalier Universitaire Vaudois  Genomics and Metagenomics Laboratory | in preparation |
| **CHUV_2101091295** | Centre Hospitalier Universitaire Vaudois  Genomics and Metagenomics Laboratory | in preparation |
| **CHUV_2101091034** | Centre Hospitalier Universitaire Vaudois  Genomics and Metagenomics Laboratory | in preparation |
| **CHUV_2101091033** | Centre Hospitalier Universitaire Vaudois  Genomics and Metagenomics Laboratory | in preparation |
| **CHUV_2101091032** | Centre Hospitalier Universitaire Vaudois  Genomics and Metagenomics Laboratory | in preparation |
| **CHUV_2101091030** | Centre Hospitalier Universitaire Vaudois  Genomics and Metagenomics Laboratory | in preparation |
| **CHUV_2101091029** | Centre Hospitalier Universitaire Vaudois  Genomics and Metagenomics Laboratory | in preparation |
| **CHUV_2101091028** | Centre Hospitalier Universitaire Vaudois  Genomics and Metagenomics Laboratory | in preparation |
| **CHUV_2101091027** | Centre Hospitalier Universitaire Vaudois  Genomics and Metagenomics Laboratory | in preparation |
| **CHUV_2101091026** | Centre Hospitalier Universitaire Vaudois  Genomics and Metagenomics Laboratory | in preparation |
| **CHUV_2101091025** | Centre Hospitalier Universitaire Vaudois  Genomics and Metagenomics Laboratory | in preparation |
| **CHUV_2101091024** | Centre Hospitalier Universitaire Vaudois  Genomics and Metagenomics Laboratory | in preparation |
| **CHUV_2101083708** | Centre Hospitalier Universitaire Vaudois  Genomics and Metagenomics Laboratory | in preparation |
| **CHUV_2101081224** | Centre Hospitalier Universitaire Vaudois  Genomics and Metagenomics Laboratory | in preparation |
| **CHUV_2101074841** | Centre Hospitalier Universitaire Vaudois  Genomics and Metagenomics Laboratory | in preparation |
| **CHUV_2101074771** | Centre Hospitalier Universitaire Vaudois  Genomics and Metagenomics Laboratory | in preparation |
| **CHUV_2101072145** | Centre Hospitalier Universitaire Vaudois  Genomics and Metagenomics Laboratory | in preparation |
| **CHUV_2101064155** | Centre Hospitalier Universitaire Vaudois  Genomics and Metagenomics Laboratory | in preparation |
| **CHUV_2101063937** | Centre Hospitalier Universitaire Vaudois  Genomics and Metagenomics Laboratory | in preparation |
| **CHUV_2101044049** | Centre Hospitalier Universitaire Vaudois  Genomics and Metagenomics Laboratory | in preparation |
| **CHUV_2101044048** | Centre Hospitalier Universitaire Vaudois  Genomics and Metagenomics Laboratory | in preparation |
| **CHUV_2101031576** | Centre Hospitalier Universitaire Vaudois  Genomics and Metagenomics Laboratory | in preparation |
| **CHUV_2012311985** | Centre Hospitalier Universitaire Vaudois  Genomics and Metagenomics Laboratory | in preparation |
| **CHUV_2012311983** | Centre Hospitalier Universitaire Vaudois  Genomics and Metagenomics Laboratory | in preparation |
| **CHUV_2012311982** | Centre Hospitalier Universitaire Vaudois  Genomics and Metagenomics Laboratory | in preparation |
| **CHUV_2012311617** | Centre Hospitalier Universitaire Vaudois  Genomics and Metagenomics Laboratory | in preparation |
| **CHUV_2012311586** | Centre Hospitalier Universitaire Vaudois  Genomics and Metagenomics Laboratory | in preparation |
| **CHUV_2012311581** | Centre Hospitalier Universitaire Vaudois  Genomics and Metagenomics Laboratory | in preparation |
| **CHUV_2012283992** | Centre Hospitalier Universitaire Vaudois  Genomics and Metagenomics Laboratory | in preparation |
| **CHUV_2012283415** | Centre Hospitalier Universitaire Vaudois  Genomics and Metagenomics Laboratory | in preparation |
| **CHUV_201226923** | Centre Hospitalier Universitaire Vaudois  Genomics and Metagenomics Laboratory | in preparation |

**Table S4.** GISAID database identifier of N501Y-carrying genomes from Switzerland included into phylogenetic analysis.
